# Benchmarking Clinical Reasoning in Large Language Models: A Comparative Assessment Study

**DOI:** 10.64898/2026.03.13.26347597

**Authors:** Thomas Prade, Matthias Samwald

## Abstract

Evaluation of Large Language Models (LLMs) and their clinical competence has mainly focused on conventional multiple-choice (MCQ) formatted medical question answering exams, yielding benchmarks like MedQA-USMLE, where models have already exceeded expert-level performance. However, alternative assessment methods have recently been proposed, such as SCT-Bench based on Script Concordance Testing (SCT), which evaluates clinical reasoning and probabilistic thinking under uncertainty. Reasoning-optimized models have unexpectedly scored worse on SCT-Bench despite outperforming non-reasoning models on other medical benchmarks. This study compared performance metrics, uncertainty proxies and clinical reasoning qualities between MedQA-USMLE and the public subset of SCT-Bench using instruction-tuned GPT-4.1, contrasting baseline and Chain-of-Thought (CoT) prompting across sampled responses. CoT prompts were designed to explicitly instruct the model to apply cognitive clinical reasoning strategies, with their usage subsequently evaluated across both benchmark formats. CoT prompting improved MedQA performance from 86.4% to 93.0%, while SCT-Bench score showed a non-significant decline from 77.7% to 74.7%. GPT-4.1 systematically overestimated the impact of new information under CoT, leading to overconfidence and increased extreme ratings on SCT questions. Sample-based majority voting significantly improved MedQA scores under CoT but had no meaningful effect on SCT-Bench. Response entropy analysis showed that CoT increased overall answer variability, while simultaneously clustering correct responses on MedQA, an effect absent on SCT-Bench. Calibration and ROC were substantially poorer on SCT-Bench than on MedQA, though CoT improved both on either benchmark. Qualitative analysis confirmed GPT-4.1 could apply situation-appropriate reasoning strategies and showed signs of metacognitive awareness about its own reasoning process, with expert rating patterns suggesting possible alignment with expert-like logic. These findings further corroborate limitations in elicited clinical reasoning for SCT-based benchmarking and suggest that reasoning-aware evaluation frameworks could contribute meaningfully to the medical AI benchmark landscape.

## 1. Introduction

Language models have since progressed to be more powerful and accurate, facilitating or taking over a range of tasks including in the biomedical domain [1]. The potential applications of LLMs in medicine are diverse, spanning direct patient care, medical education, and research [2]. Since these tasks require domain-specific competence, biomedical language models such as BioMedLM, BioGPT, Clinical Camel, and MedAlpaca have been introduced [1], and have shown enhanced performance on clinical text mining, biomedical information extraction and medical question answering compared to general-purpose models [1]. LLMs can enrich biomedical queries [1] and have shown higher precision than state-of-the-art methods when formulating boolean queries for systematic reviews [3]. Biomedical text summarization has proven useful for medical literature, radiology reports and clinical notes, though challenges remain in maintaining clinical accuracy and avoiding omission of crucial information [1]. Information extraction from unstructured clinical notes can help present patient data in structured form, contributing to CDS [4]. In clinical workflows, LLMs could reduce administrative burden, such as report writing, freeing physicians for patient care and interdisciplinary collaboration [2]. In medical education, LLMs show potential as interactive tools through case generation, follow-up questioning and counterfactual reasoning to address student misconceptions [1, 2]. However, implementing AI-powered CDSSs raises significant concerns [4]: systematic biases from historical data risk discriminatory recommendations; interoperability with existing healthcare IT remains technically challenging; and physicians express valid concerns about clinical accuracy and legal liability. Data security, confidentiality and broader ethical considerations further complicate deployment. The highlighted potential use-cases offer great opportunities for optimizing healthcare workflow, particularly via human-AI hybrid systems [4, 5]. For developing sophisticated CDSSs, it is fundamental that LLMs are capable to correctly process medical data and use the information adequately when performing different kind of tasks. This includes e.g. summarising patient records [1], generating content while avoiding hallucinations [2] or assisting triage by identifying time-sensitive conditions [6]. Performing these tasks requires clinical reasoning: correct pathophysiological inference, adequate use of medical knowledge, and metacognitive awareness of reasoning strategies fundamental to medical practice [7, ch. 1][8][7, ch. 4][9, 10].

### 1.1. Clinical Reasoning

Clinical reasoning is a complex cognitive process fundamental to medical practice, involving diverse thinking strategies operating through both conscious and unconscious mechanisms [9, 11]. Its core components include information gathering, hypothesis generation, forming a problem representation, generating a differential diagnosis, and developing a management plan [11].

Knowledge alone is insufficient for both future doctors and AI systems. Internalizing the principles of clinical thinking is essential, as is having effective ways to evaluate whether they are capable of doing so. Emotions also play a role in human clinical decision making, particularly in high-stakes situations where they interact with cognitive processes [12], an element almost certainly absent in AI systems.

Errors in reasoning play a significant role in diagnostic and therapeutic mistakes that may compromise patient safety [7, pp. 20-21]. Framing clinical reasoning within dual-process theory has shown that both System 1 and System 2 are equally prone to errors, albeit for different reasons [10]. Cooper and Frain categorized important error sources as follows [7, pp. 20-21]: (1) no-fault errors (unavoidable errors due to missing or withheld information), (2) system errors (organizational and contextual factors such as lack of resources, deficient supervision, or communication failures), (3) knowledge gaps (where even ideal reasoning cannot compensate for substantial deficits), (4) misinterpretation of diagnostic tests and their effect on disease probabilities, and (5) cognitive biases. The question to what extent LLMs can fall into the pitfalls of such biases or tend to replicate them in the context of clinical decision making remains largely unexplored.

LLMs and other AI systems possess great potential to excel in the theoretical frameworks that describe clinical reasoning, though human experts arguably retain performance advantages in real-world settings due to decades of rigorous clinical experience [13]. Motivated by cognitive processes involved in clinical decision making [7, ch. 4], Table 1 provides an overview of a set of reasoning strategies and their use cases. Many of these thinking schemes are interdependent and consist of elements from one another, and the table can be understood as a simplified toolkit for clinical reasoning that was integrated in the prompts used for this study (section 3).

**Table 1:**
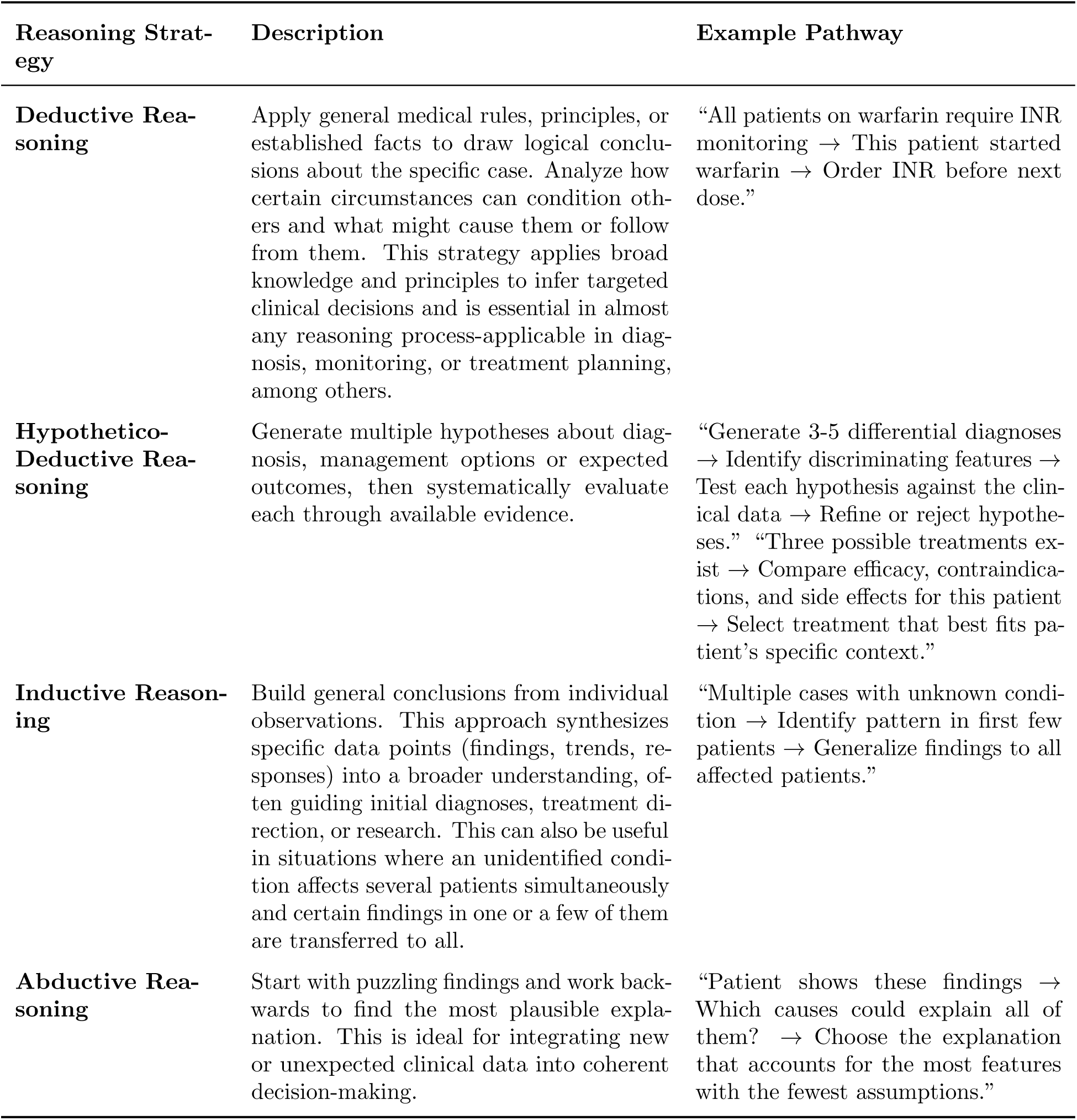
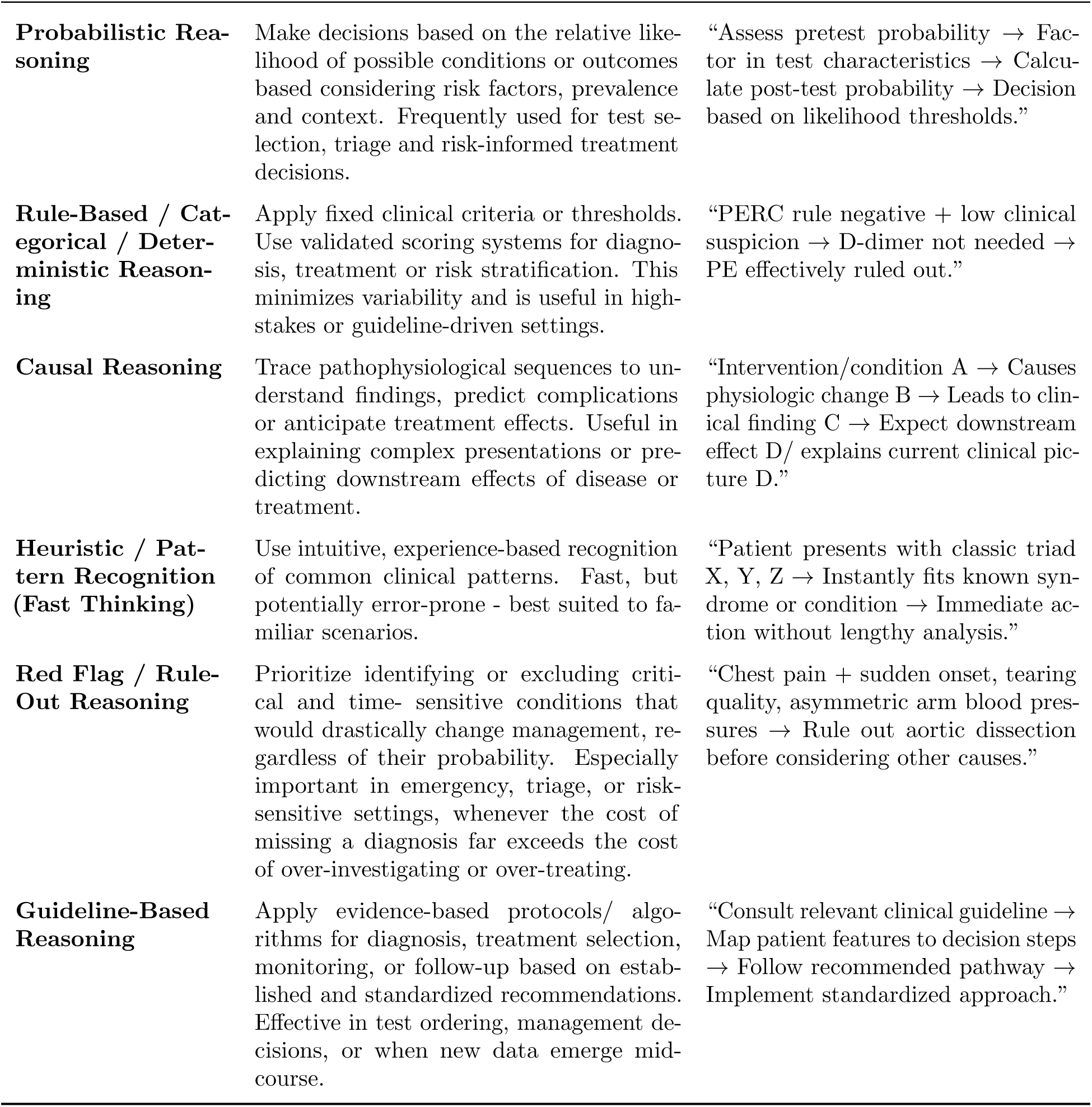
Overview of clinical reasoning strategies with brief descriptions and illustrative pathways. Own representation based on strategy mentions in the literature [7, ch. 4][9].

| Reasoning Strat-egy | Description | Example Pathway |
|---|---|---|
| **Deductive Rea-soning** | Apply general medical rules, principles, or established facts to draw logical conclu-sions about the specific case. Analyze how certain circumstances can condition oth-ers and what might cause them or follow from them. This strategy applies broad knowledge and principles to infer targeted clinical decisions and is essential in almost any reasoning process-applicable in diag-nosis, monitoring, or treatment planning, among others. | “All patients on warfarin require INR monitoring → This patient started warfarin → Order INR before next dose.” |
| **Hypothetico-Deductive Rea-soning** | Generate multiple hypotheses about diag-nosis, management options or expected outcomes, then systematically evaluate each through available evidence. | “Generate 3-5 differential diagnoses → Identify discriminating features → Test each hypothesis against the clin-ical data → Refine or reject hypothe-ses.” “Three possible treatments ex-ist → Compare efficacy, contraindica-tions, and side effects for this patient → Select treatment that best fits pa-tient’s specific context.” |
| **Inductive Reason-ing** | Build general conclusions from individual observations. This approach synthesizes specific data points (findings, trends, re-sponses) into a broader understanding, of-ten guiding initial diagnoses, treatment di-rection, or research. This can also be useful in situations where an unidentified condi-tion affects several patients simultaneously and certain findings in one or a few of them are transferred to all. | “Multiple cases with unknown condi-tion → Identify pattern in first few patients → Generalize findings to all affected patients.” |
| **Abductive Rea-soning** | Start with puzzling findings and work back-wards to find the most plausible expla-nation. This is ideal for integrating new or unexpected clinical data into coherent decision-making. | “Patient shows these findings → Which causes could explain all of them? → Choose the explanation that accounts for the most features with the fewest assumptions.” |
| **Probabilistic Rea-soning** | Make decisions based on the relative like-lihood of possible conditions or outcomes based considering risk factors, prevalence and context. Frequently used for test se-lection, triage and risk-informed treatment decisions. | “Assess pretest probability → Fac-tor in test characteristics → Calcu-late post-test probability → Decision based on likelihood thresholds.” |
| **Rule-Based / Cat-egorical / Deter-ministic Reason-ing** | Apply fixed clinical criteria or thresholds. Use validated scoring systems for diagno-sis, treatment or risk stratification. This minimizes variability and is useful in high-stakes or guideline-driven settings. | “PERC rule negative + low clinical suspicion → D-dimer not needed → PE effectively ruled out.” |
| **Causal Reasoning** | Trace pathophysiological sequences to un-derstand findings, predict complications or anticipate treatment effects. Useful in explaining complex presentations or pre-dicting downstream effects of disease or treatment. | “Intervention/condition A → Causes physiologic change B → Leads to clin-ical finding C → Expect downstream effect D/ explains current clinical pic-ture D.” |
| **Heuristic / Pat-tern Recognition (Fast Thinking)** | Use intuitive, experience-based recognition of common clinical patterns. Fast, but potentially error-prone - best suited to fa-miliar scenarios. | “Patient presents with classic triad X, Y, Z → Instantly fits known syn-drome or condition → Immediate ac-tion without lengthy analysis.” |
| **Red Flag / Rule-Out Reasoning** | Prioritize identifying or excluding criti-cal and time- sensitive conditions that would drastically change management, re-gardless of their probability. Especially important in emergency, triage, or risk-sensitive settings, whenever the cost of missing a diagnosis far exceeds the cost of over-investigating or over-treating. | “Chest pain + sudden onset, tearing quality, asymmetric arm blood pres-sures → Rule out aortic dissection before considering other causes.” |
| **Guideline-Based Reasoning** | Apply evidence-based protocols/ algo-rithms for diagnosis, treatment selection, monitoring, or follow-up based on estab-lished and standardized recommendations. Effective in test ordering, management de-cisions, or when new data emerge mid-course. | “Consult relevant clinical guideline → Map patient features to decision steps → Follow recommended pathway → Implement standardized approach.” |

**Table 2:**
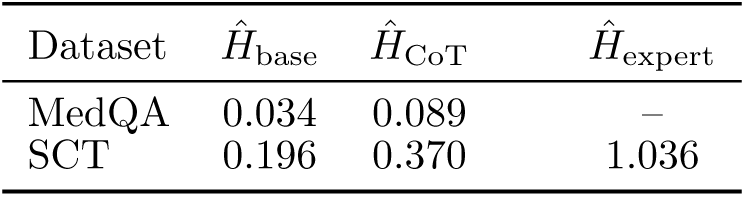
Mean entropy values depending on dataset and prompting mode.

| Dataset | Ĥ<sub>base</sub> | Ĥ<sub>CoT</sub> | Ĥ<sub>expert</sub> |
|---|---|---|---|
| MedQA | 0.034 | 0.089 | – |
| SCT | 0.196 | 0.370 | 1.036 |

### 1.2. Clinical Prompt Engineering

Several studies have examined strategies for enhancing clinical reasoning in LLMs with varying success using different prompt-engineering techniques [14, 15, 16, 17, 18, 19]. Chain-of-thought (CoT) prompting has shown how making reasoning processes explicit through intermediate steps can enhance LLM performance on complex tasks [20]. However, the effectiveness of CoT for medical problem solving varies depending on the task and context [15].

A common denominator across approaches in the literature is imitating human cognitive processes in clinical decision making, though effectiveness varies significantly across assessment formats, particularly under uncertainty. McCoy et al. introduced SCT as a novel benchmarking approach for LLM clinical reasoning [21], finding that LLMs match or exceed medical student performance but consistently lag behind senior residents and expert physicians. Most notably, reasoning models unexpectedly underperformed on SCT-Bench compared to their scores on previous medical benchmarks, leading the authors to claim it as the first medical benchmark where enhanced reasoning decreases rather than improves performance [21]. This suggests that SCTs evaluate reasoning skills beyond encoded biomedical knowledge and may be able to unmask LLM limitations in clinical judgement [21].

### 1.3. Medical Benchmarking

The evaluation of LLMs in medical contexts has been anchored in part by traditional medical licensing exams like the United States Medical Licensing Examination (USMLE) [2, 14, 22]. The most recent general purpose models have reached superhuman levels, with OpenAI’s reasoning model o1 currently leading the MedQA leaderboard at 96.5%, approaching the practical ceiling of the benchmark [23]. These outstanding performances have generated considerable optimism about future clinical applications. However, traditional medical benchmarks carry significant limitations that prevent translating LLM accuracy to real-world performance [21, 13].

High scores on USMLE-style datasets likely reflect encoded knowledge and test-taking ability rather than genuine clinical reasoning [21]. Moreover, multiple-choice formats do not adequately represent the complexity of real clinical decision making, which involves collecting information from multiple sources, iterative re-evaluation, and continuous adaptation across decision steps, all within a highly interactive and uncertain environment [13]. In contrast, LLMs have been tested primarily in structurally simple contexts such as case vignettes with all information provided upfront [13]. Concerns about data contamination and memorization further compound these limitations [2]. Acknowledging the limitations certain assessment methods have has led to recognizing the necessity for alternative and more sophisticated evaluation frameworks.

### 1.4. Script Concordance Testing

SCTs have been designed to evaluate clinical reasoning under uncertainty, a fundamental skill hardly assessed by common MCQ exams [21]. SCT is grounded in script theory from cognitive psychology, which proposes that clinicians use organized knowledge structures (scripts) to make sense of clinical problems and guide decisions, with these scripts becoming more refined through clinical experience [24]. Rather than testing recall of medical facts, SCT focuses on how clinicians integrate new information to modify their reasoning under ambiguity [25]. Instead of presenting precise, complete scenarios with single correct answers, SCT presents situations where new information must be weighed against existing hypotheses, mimicking real-life aspects of clinical decision making [25].

### 1.5. Uncertainty Quantification

Accurately quantifying and communicating uncertainty is a fundamental requirement for deploying LLMs in clinical settings, where overconfident incorrect predictions could have life-threatening consequences. Calibration, meaning how well a model’s confidence matches its actual accuracy, is especially important in medicine, as reliable probability estimates would help clinicians relativize outputs accordingly [2]. However, the inability of LLMs to reliably communicate uncertainty remains a significant barrier to clinical implementation [26]. Uncertainty quantification methods broadly fall into white-box approaches requiring internal model access and black-box methods using only model outputs [27]. Black-box methods are essential for closed-source commercial APIs and include sample-based and self-evaluation approaches of varying sophistication [27].

Confidence elicitation consistently demonstrates systematic overconfidence [26, 28], though this contrasts with studies concluding that LLMs mostly “know what they know” [29]. Liévin et al. found that models such as Llama 2 were well calibrated on medical benchmarks when using 5-shot CoT prompting [30], and Steyvers et al. suggest the underlying problem lies in how confidence is communicated rather than in poor model self-awareness [31].

Confidence scores and uncertainty parameters are necessary for estimating both calibration and discriminative ability between accurate and inaccurate predictions. Savage et al. evaluated confidence elicitation, token-level probability, and sample consistency metrics across several models including GPT-3.5 and GPT-4 for medical diagnosis and treatment tasks, finding that sample consistency was generally the superior method and that uncertainty assessment was task-specific, with almost all metrics showing better discriminative ability for diagnosis than treatment questions [26].

## 2. Methods

### 2.1. Datasets

The primary sources for medical test questions and case vignettes were two datasets: MedQA-USMLE [32], extensively used for benchmarking LLMs in biomedicine, and SCT-Bench introduced by McCoy et al. [21]. Both benchmarks evaluate clinical competence, yet assess different qualities and aspects of it.

#### 2.1.1. MedQA-USMLE

MedQA served as both the primary reference for conventional MCQ assessment and the comparison point for SCT. A subset of 700 USMLE questions spanning steps 1 to 3 was used to evaluate GPT-4.1 [33]. Each item presents 5 answer options with one correct answer, and the final score is the percentage of correctly answered questions.

#### 2.1.2. SCT-Bench

McCoy et al. published SCT-Bench comprising 750 questions across 10 SCT datasets from diverse medical specialties originating in North America, Asia, and Australia [21]. Only a smaller public subset of 174 questions was used here, combining Open Medical SCT (University of Florida, 72 paediatrics questions) and Adelaide SCT (University of Adelaide, 102 multi-specialty questions). The expert consensus is scored at 1.0, with other ratings weighted as fractions based on the proportion of experts selecting each option, and responses chosen by no expert receiving 0 points [21].

### 2.2. Model and Data Generation

This study used OpenAI’s GPT-4.1 (gpt-4.1-2025-04-14). Unlike reasoning-optimized models such as the o-series, GPT-4.1 lacks built-in CoT features and relies on explicit instructional prompting. OpenAI notes its particular suitability for tasks requiring precise adherence to specified formats and multi-step procedures [33], making it a good candidate for studying the impact of CoT-inducing prompts compared to a zero-instruction baseline. Temperature was set to 0.7 and top-p to 1.0 for all queries. Prompts were sent via OpenAI API to GPT-4.1, with responses written to structured JSON files for subsequent analysis, including extracted answers, confidence scores, and reasoning strategy data. The publicly available query code from McCoy et al. [21] served as a template and was adapted for this study. AI assistance was used for certain text extraction and data analysis functions [34, 35]. Code and data availability information can be found in the supplementary materials (Appendix C).

### 2.3. Prompting and Uncertainty Quantification

Two types of data queries were run:

i. <u>Baseline prompts with no CoT:</u> Zero-instruction prompts directing the model to answer without reasoning steps or CoT.
ii. <u>Clinical CoT prompts:</u> Prompts augmented with instructions for clinical CoT, incremental information processing and a list of clinical reasoning strategies based on Table 1. Prompt templates can be found in Appendix B.

In every prompt, GPT-4.1 was instructed to rate its confidence in its response on a scale from 0 to 100, where 0 represents complete uncertainty and 100 complete certainty. This one-step confidence elicitation approach is not highly accurate, but was employed for its simplicity and negligible additional cost. Inspired by Savage et al. [26], uncertainty was additionally quantified via sample consistency. Each question was sent 15 times (n = 15) to the model, a sample size at which consistency performance has generally reached a plateau [36]. The most frequent response was taken as the reference answer for determining correctness, and the sample consistency score was calculated as the fraction of responses matching this majority response (X/n). Wang et al. showed that combining multiple sampled reasoning paths with majority voting improves accuracy significantly over single CoT prompting [37]. For CoT prompts, reasoning strategy extraction was additionally implemented: the model was instructed to identify and report which clinical reasoning strategies it applied from a predefined list (see Table 1), enabling analysis of strategy distribution and its relationship to performance outcomes.

#### 2.3.1. ROC Curve

The receiver operating characteristic (ROC) curve [38] evaluates classification performance by plotting the true positive rate (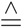 = sensitivity) against the false positive rate (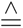 = 1*−*specificity) across all decision thresholds [39]. At each threshold *τ*, predictions with confidence *≥ τ* are classified as correct, yielding:

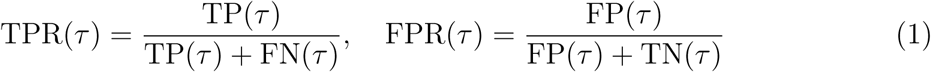

where TP, FN, FP, and TN denote true positives, false negatives, false positives, and true negatives respectively.

#### 2.3.2. Expected Calibration Error

The expected calibration error (ECE) quantifies the alignment between predicted confidence and actual accuracy, measuring how well calibrated a model is [40]. A model is well calibrated if an outcome predicted with probability *p* occurs approximately *p* fraction of the time across the full predicted range [40]. ECE is computed by partitioning predictions into *M* equal-width bins and taking the weighted average of absolute differences between mean confidence and accuracy per bin [31]:

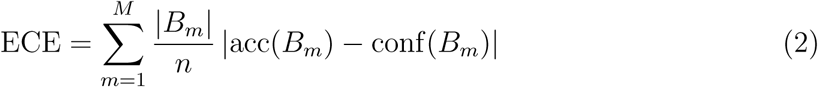

where *B_m_*is the *m*-th bin, *n* is the total number of predictions, acc(*B_m_*) is the fraction of correct predictions in bin *m*, and conf(*B_m_*) is the mean predicted confidence in that bin.

### 2.4. Evaluation Metrics and Data Analysis

The primary evaluation metric for both benchmarks is the test score, calculated in two ways: using majority responses as the reference answer, and by randomly sampling answers to derive an average score for comparison with the majority score. Further metrics include response entropy analysis, ROC and calibration parameters, quantitative and qualitative analysis of self-reported reasoning strategy use and LLM rating distributions as well as CoT-influenced rating transitions on SCT-Bench. Statistical testing for benchmark scores and response entropies was performed using paired and unpaired t-tests as well as McNemar’s test.

## 3. Results

### 3.1. Majority Vote

The majority MedQA score at baseline prompting is 86.4%, while clinical CoT prompting achieved 93%, representing a statistically significant improvement of almost 7 percentage points (*p <* 0.05) (Figure 1a).

**Figure 1:**
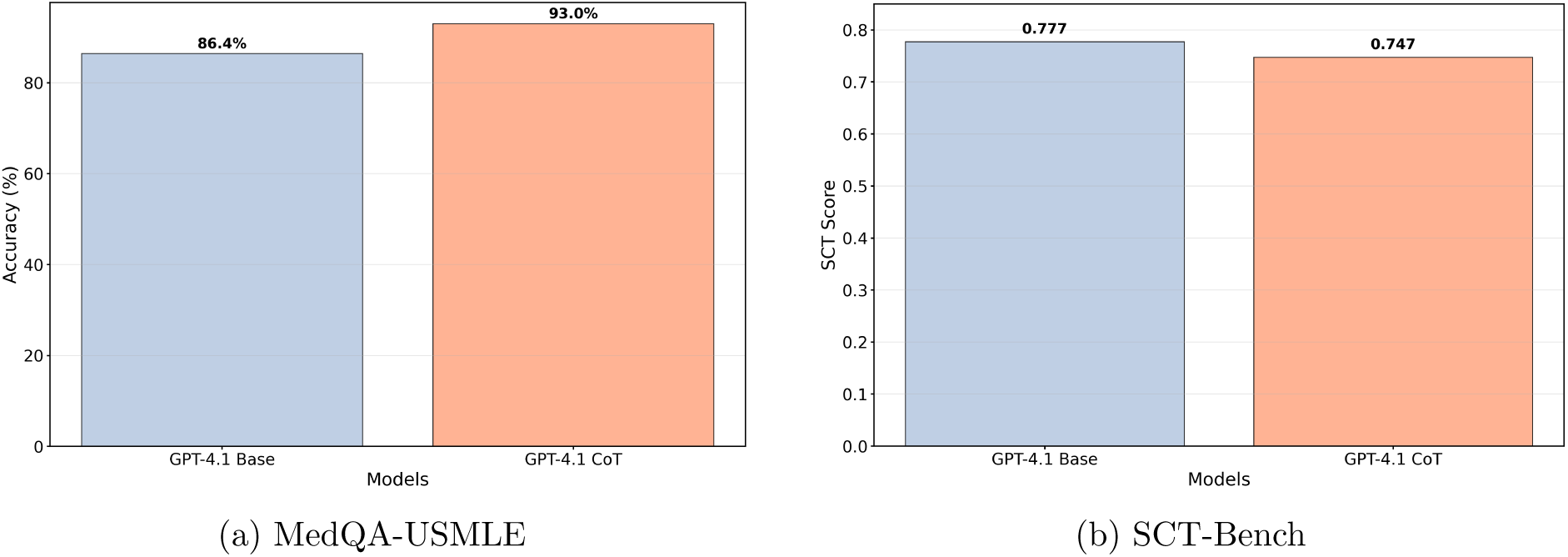
Majority benchmark scores with vs. without clinical CoT prompting.

Results for SCT-Bench are not consistent with those for MedQA. The majority SCT-Bench score at baseline is 77.7%, dropping to 74.7% with CoT prompting (Figure 1), though this decrease of 3 percentage points was not statistically significant (*p ≈* 0.45).

### 3.2. Mean Score

The mean MedQA score also improved by approximately 5 percentage points (*p <* 0.05) (Figure 2). At baseline, the majority and mean scores differ by only 0.1 percentage points, compared to 1.4 percentage points under CoT. Notably, sample-based majority voting significantly benefited performance only under CoT (*p <* 0.05), while for baseline prompting it offered no significant advantage over a single response (*p* ≈ 0.96).

**Figure 2:**
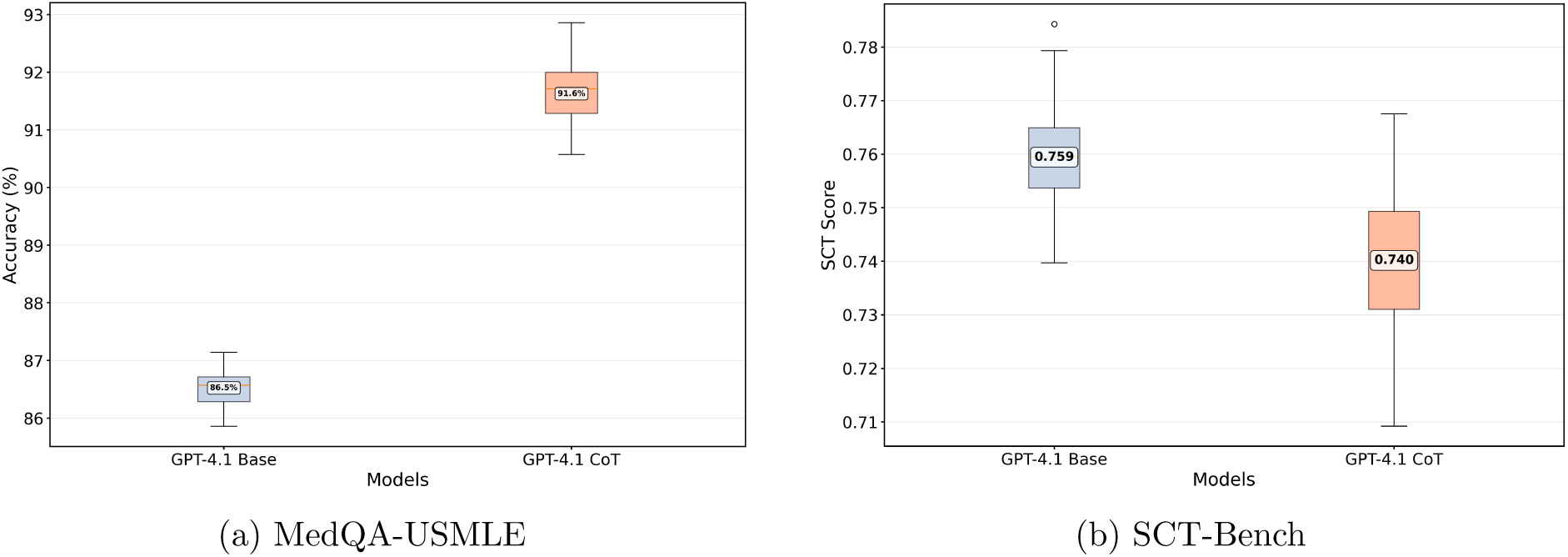
Mean benchmark scores (N = 100) with vs. without clinical CoT prompting.

The mean SCT score dropped from 75.9% to 73.8%, which was not statistically significant (*p ≈* 0.54). Majority voting marginally improved scores in both prompting modes (*p <* 0.05), though the benefit was greater for baseline than CoT, reflecting reduced consistency of correct answers under CoT on SCT-Bench.

Overall, clinical CoT prompting had a positive impact only on MedQA, increasing benchmark scores and the frequency of correct answers to a degree that made sampling and majority voting significantly beneficial.

### 3.3. Response Entropy

Response entropy quantifies disagreement among LLM responses to the same question. On MedQA, mean entropy increased significantly with clinical CoT (*p <* 0.05) from *Ĥbase* = 0.034 to *ĤCoT* = 0.089 (Figure 3a), indicating greater response variability. Notably, despite this increased dispersion, the correct answer became more likely to be selected by majority voting, indicating stronger clustering around the correct response.

**Figure 3:**
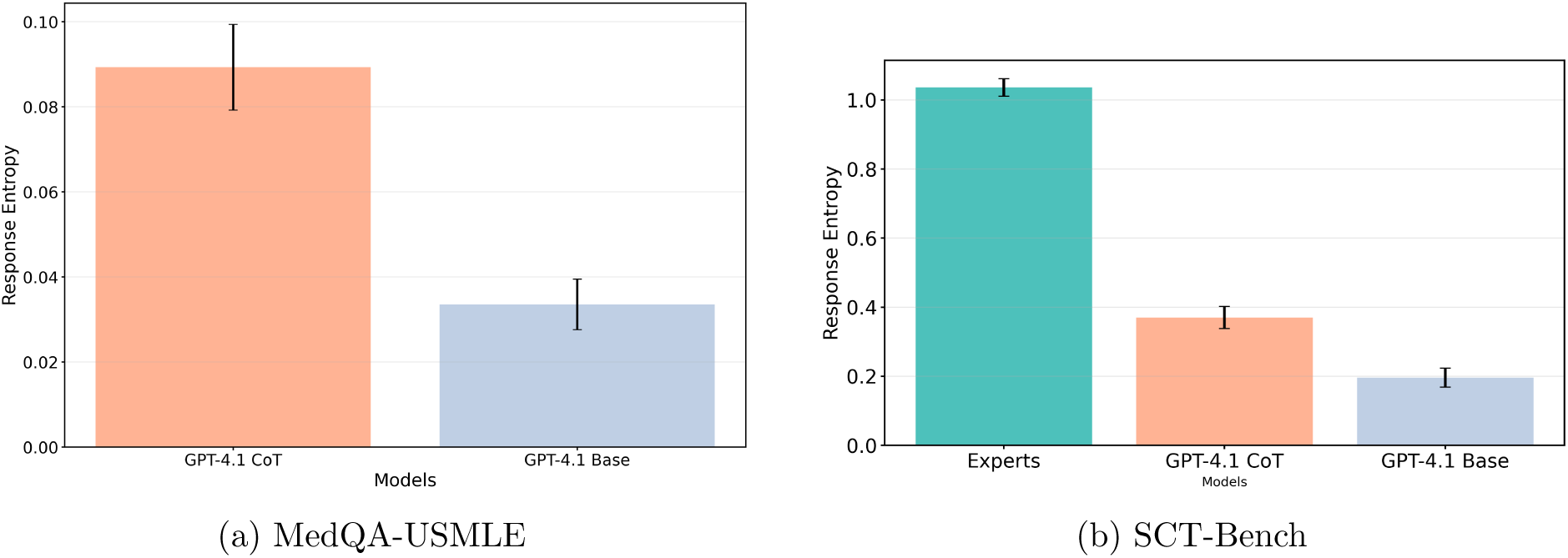
GPT-4.1 response entropy on MedQA-USMLE and SCT-Bench with vs. without clinical CoT prompting.

Clinical CoT had the same effect on rating variability on SCT-Bench (Figure 3b). However, unlike on MedQA, increased variability did not increase the frequency of correct answers but rather reduced it. Since SCT is based on multiple reference expert answers, non-zero variance among expert ratings is also present.

As shown in Figure 3b, expert response entropy is considerably higher than model entropy. While most experts tend to agree, dissenting opinions are common. Clinical CoT significantly increased GPT-4.1’s response entropy compared to baseline (*p <* 0.05), but without the cluster formation around correct answers observed on MedQA.

### 3.4. Calibration Metrics

Calibration metrics were computed using both self-elicited confidence and sample consistency as described in subsection 2.3. Self-elicited confidence proved ineffective as a calibration signal. GPT-4.1 reported confidence scores only within a very limited range, rarely below 60%, making complete calibration curves uninterpretable. The model was systematically overconfident on both datasets. Clinical CoT had minimal impact on confidence elicitation but reduced overconfidence slightly on MedQA, decreasing ECE by more than half. On SCT-Bench, CoT had almost no effect on elicitation quality and only little positive impact on ECE.

#### 3.4.1. Sample Consistency

Sample consistency provided a broader uncertainty range than self-elicited confidence. On MedQA, calibration improved considerably Figure 4. This magnitude of impact was not reproducible on SCT-Bench, where calibration was substantially worse overall and CoT improved it only minimally. An overview of all ECE values is provided in Table 3.

**Figure 4:**
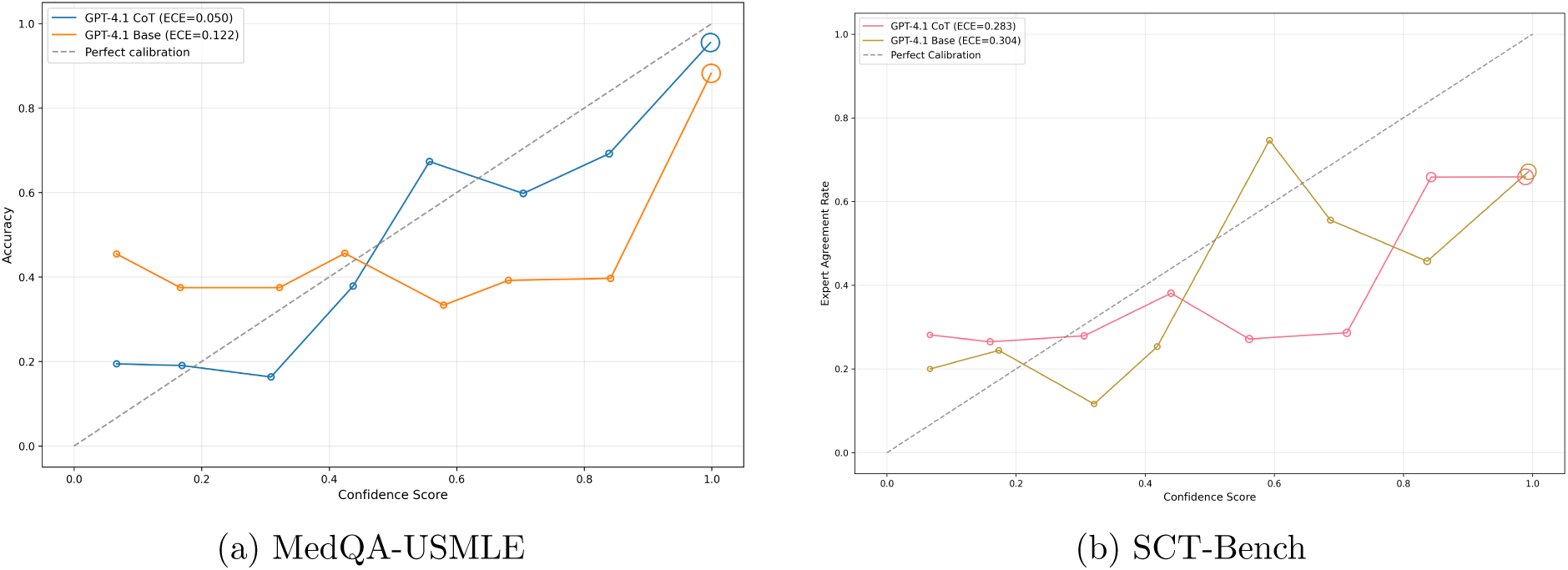
Calibration curves for sample-based uncertainty estimation with and without clinical CoT prompting. Circle radius correlates with data occupancy of each bin.

**Table 3:** Rounded AUROC and ECE values on MedQA-USMLE and SCT-Bench depending on uncertainty estimation method and prompting mode.

| **Benchmark** | **Uncertainty Proxy** | **Prompting** | **AUROC** | **ECE** |
|---|---|---|---|---|
| MedQA | Self-elicited | Baseline | 0.73 | 0.1 |
|  |  | CoT | 0.84 | 0.05 |
|  | Sample-based | Baseline | 0.61 | 0.13 |
|  |  | CoT | 0.71 | 0.05 |
| SCT-Bench | Self-elicited | Baseline | 0.6 | 0.31 |
|  |  | CoT | 0.66 | 0.31 |
|  | Sample-based | Baseline | 0.53 | 0.3 |
|  |  | CoT | 0.66 | 0.28 |

#### 3.4.2. ROC

Clinical CoT consistently increased AUROC on both datasets and both uncertainty proxies (subsubsection 2.3.1). On MedQA, AUROC improved by 14.7 percentage points using self-elicited confidence (Figure 5a) and by 17.5 percentage points using sample consistency (Figure 6a), with discrimination particularly enhanced in the intermediate confidence range. On SCT-Bench, CoT improved AUROC by 10.7 percentage points with self-elicited confidence and by 23.3 percentage points with sample-based uncertainty (Figure 5b, Figure 6b). At baseline, SCT-Bench sample-based discrimination was barely above chance. Overall, both calibration and ROC performance were substantially better on MedQA than SCT-Bench (Table 3).

**Figure 5:**
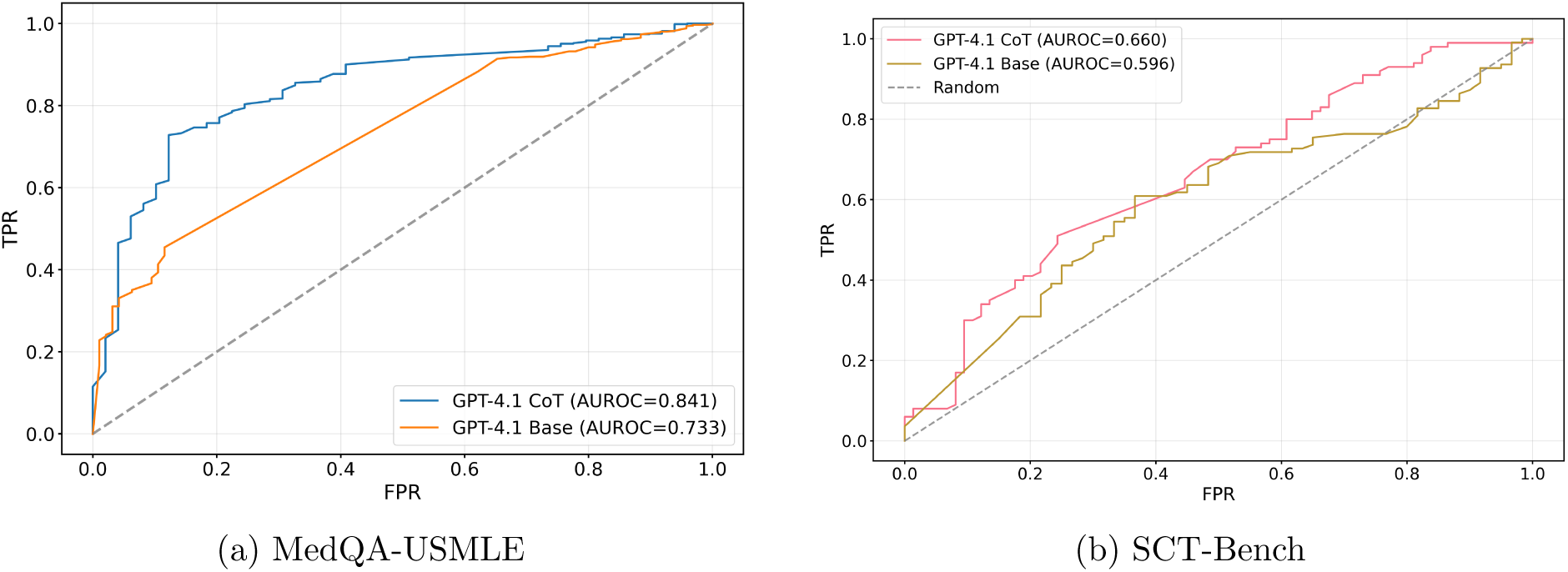
ROC curves for one-step self-elicited confidence with and without clinical CoT prompting.

**Figure 6:**
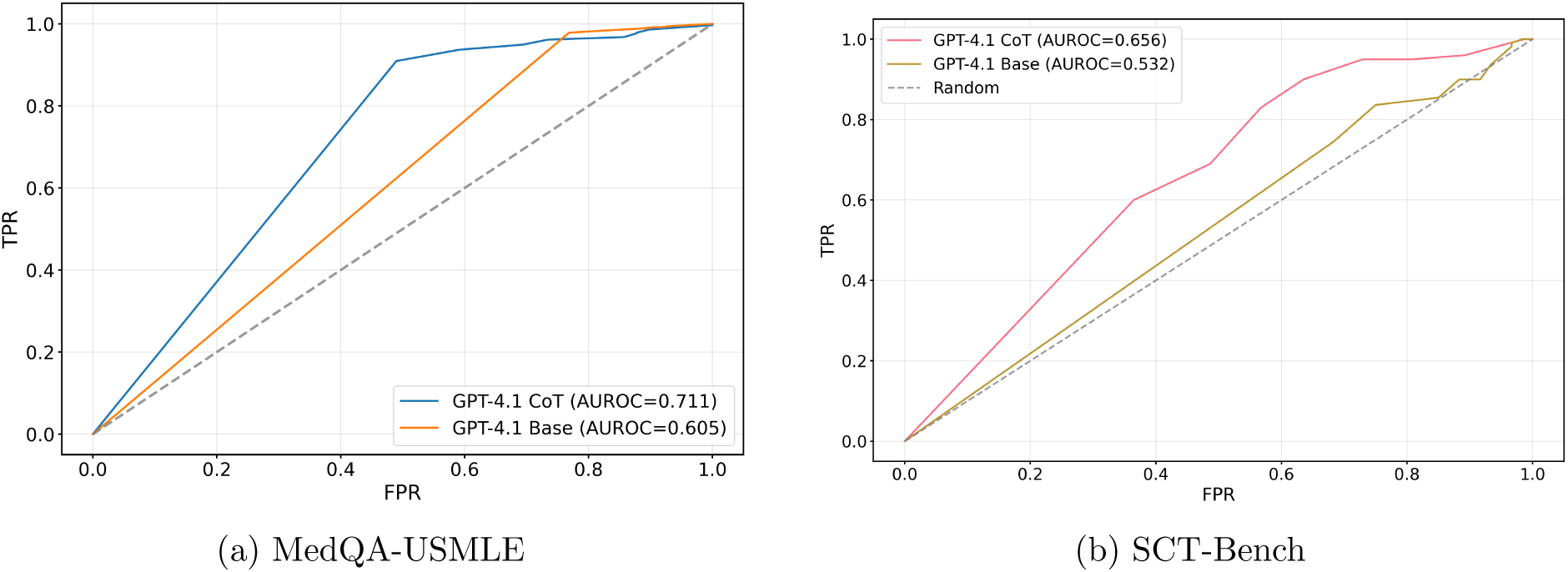
ROC curves for sample-based confidence estimation with and without clinical CoT prompting.

### 3.5. SCT Rating Distribution

The SCT score measures agreement with expert consensus, where a perfect score of 1.0 implies full alignment. However, deviating from consensus does not necessarily indicate clinical incompetence, as SCT inherently involves uncertainty and expert rating variability. We define a rating disagreement parameter Δ*r*:

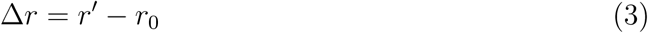

where *r^′^*is any chosen rating and *r*_0_ is the consensus expert rating.

At baseline, the Jensen-Shannon distance between expert and GPT-4.1 rating distributions is 0.109, indicating moderate similarity (Figure 7a). The model showed a slightly greater tendency toward strong disagreement (|Δ*r| >* 1) compared to physicians, who rarely deviated by more than *|±*1|. Clinical CoT increased this tendency markedly, with the model more frequently selecting extreme ratings and the Jensen-Shannon distance rising to 0.127 (Figure 7b).

**Figure 7:**
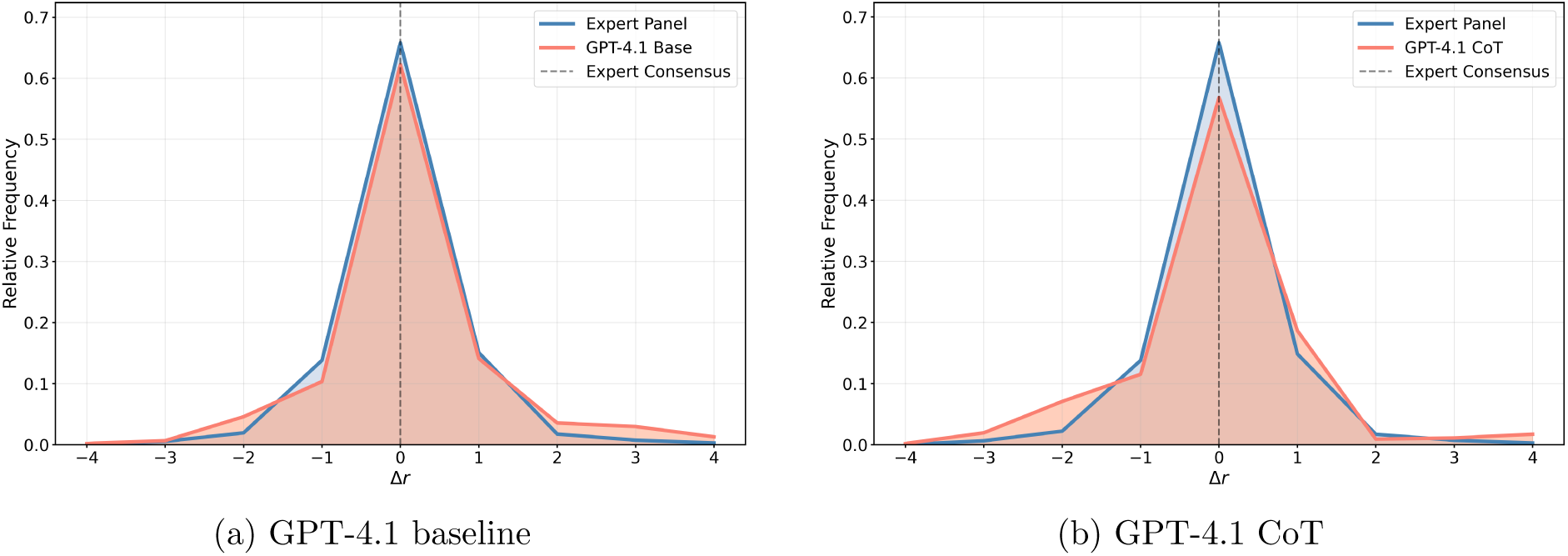
SCT-Bench rating disagreement distributions around the centered expert consensus rating at baseline and with CoT prompting.

### 3.6. Clinical Reasoning Strategies

For each dataset, GPT-4.1 was instructed to apply clinical reasoning strategies from Table 1 alongside CoT prompting, with the goal of assessing whether explicitly emulating clinical thinking would measurably affect benchmark scores and whether the model would effectively implement these strategies in its chain-of-thought.

A priori, self-reported reasoning strategies cannot be assumed to align with actual CoT output. LLMs do not engage in genuine cognitive processes, but can mirror human thought patterns. Evaluating this alignment requires manual inspection of whether the illustrated reasoning paths genuinely depict the reported strategies. Generally, the self-reported strategies were quite consistent with the output CoT steps and rationale (see Appendix A).

Self-reporting of reasoning strategies is inherently susceptible to errors: some strategies may go unreported despite being applied, while others may be mentioned without genuine implementation. Although the extent of these limitations could not be fully assessed within the scope of this study, the model demonstrably emulated clinical reasoning processes and showed signs of meta-cognitive “awareness” of doing so. With these caveats in mind, the self-reported strategy data remain insightful.

#### 3.6.1. Strategy Distribution

Examining the distribution of self-reported reasoning strategies reveals that certain schemes were reported significantly more frequently than others. Figure 8a shows the strategy histogram for MedQA, where three dominate: deductive, guideline-based, and hypothetico-deductive reasoning. Deductive reasoning is a core element of virtually any problem-solving process. Hypothetico-deductive reasoning is closely related but starts from specific clinical hypotheses such as a differential diagnosis, making it naturally prominent in a multiple-choice format where answer options already represent hypotheses to iterate through. Guideline-based reasoning is pivotal in real clinical practice and was correspondingly frequent in GPT-4.1’s outputs.

**Figure 8:**
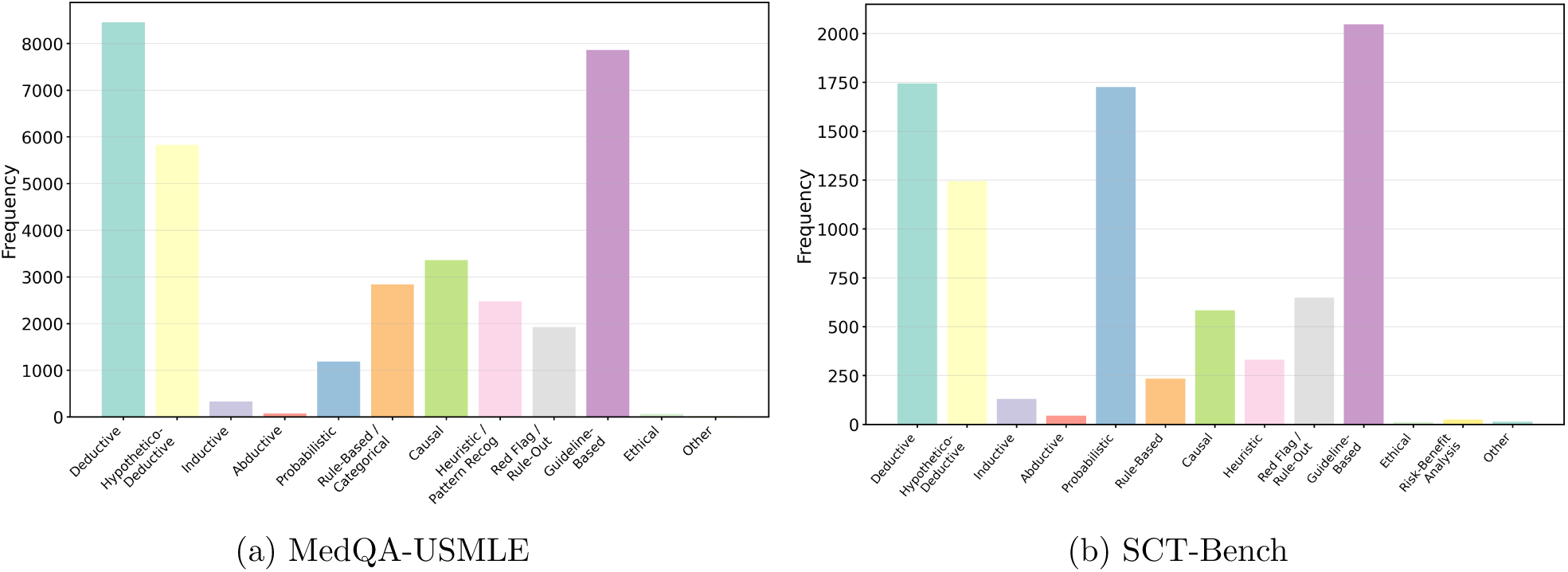
Distribution of self-reported clinical reasoning strategies.

Inductive and abductive reasoning were rarely reported. Abductive reasoning is most naturally applied when generating hypotheses such as a differential diagnosis [7, p. 18], but since MCQ formats already supply hypotheses as answer options, the model consistently favored a hypothetico-deductive approach instead. Similarly, inductive reasoning, which involves generalizing from specific observations [7, p. 18], had little applicability in this context.

Compared to SCT-Bench (Figure 8b), rule-based and heuristic reasoning were relatively more represented on MedQA, which may reflect the nature of the two formats: MedQA vignettes provide rich clinical pictures that lend themselves to pattern recognition, while SCT’s focus on uncertainty may reduce the applicability of such categorical strategies.

The strategy distribution for SCT-Bench (Figure 8b) reveals a key difference from MedQA: probabilistic reasoning ranks among the top three strategies, consistent with SCT’s focus on likelihood shifts and reasoning under uncertainty.

### 3.7. Rating Transitions

Comparing Figure 9a and Figure 9b reveals the effect of CoT on GPT-4.1’s rating behavior: strong ratings (+2, *−*2) become more frequent while moderate and neutral ratings decrease, with the exception of *r* = *−*1, which increases slightly due to unexpected flipping from *r* = +1.

**Figure 9:**
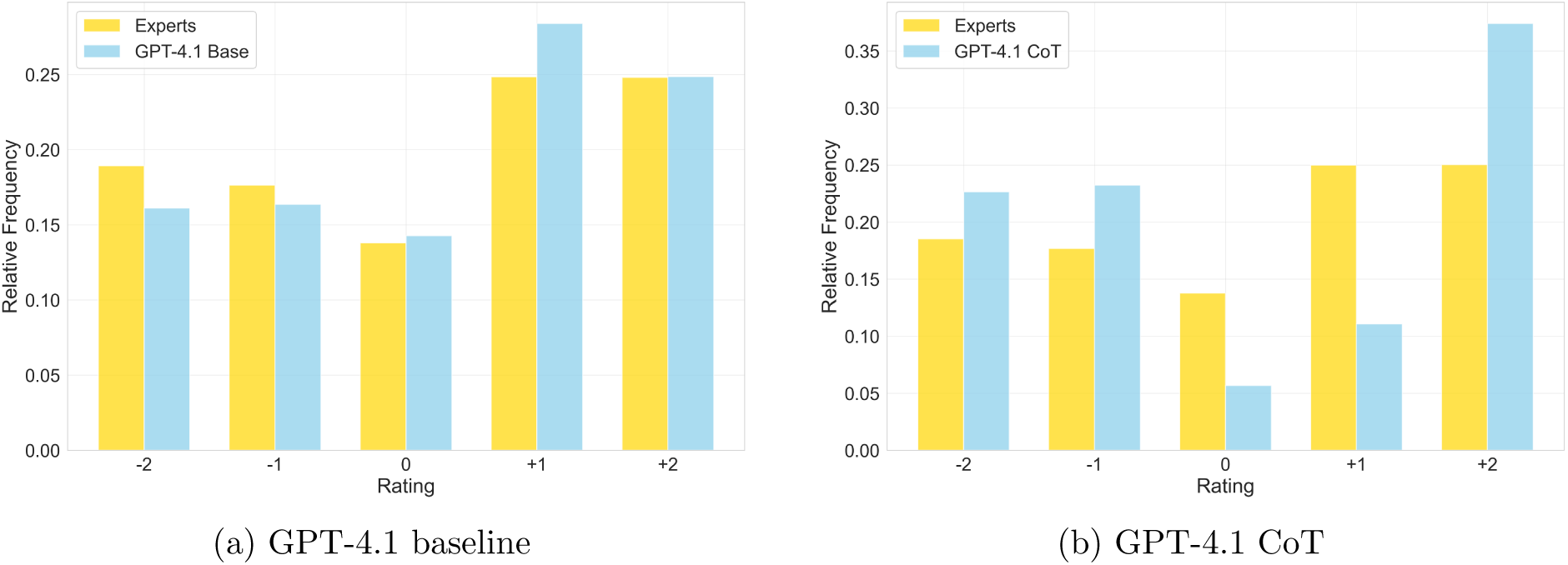
Distribution of expert and GPT-4.1 ratings at baseline and with CoT prompting.

However, the histograms cannot capture how individual SCT items were affected. Sankey diagrams were therefore used to visualize rating transitions within the same questions across raters and prompting modes, with ratings grouped into three categories: Neutral, Moderate and Strong.

Under CoT, the model shows a tendency to assign stronger ratings than experts do. Neutral ratings transition mostly to moderate ones. Overall, ratings shift systematically from weaker to stronger categories, an effect exposed explicitly in Figure 10b, which shows transitions within GPT-4.1 itself between baseline and CoT.

**Figure 10:**
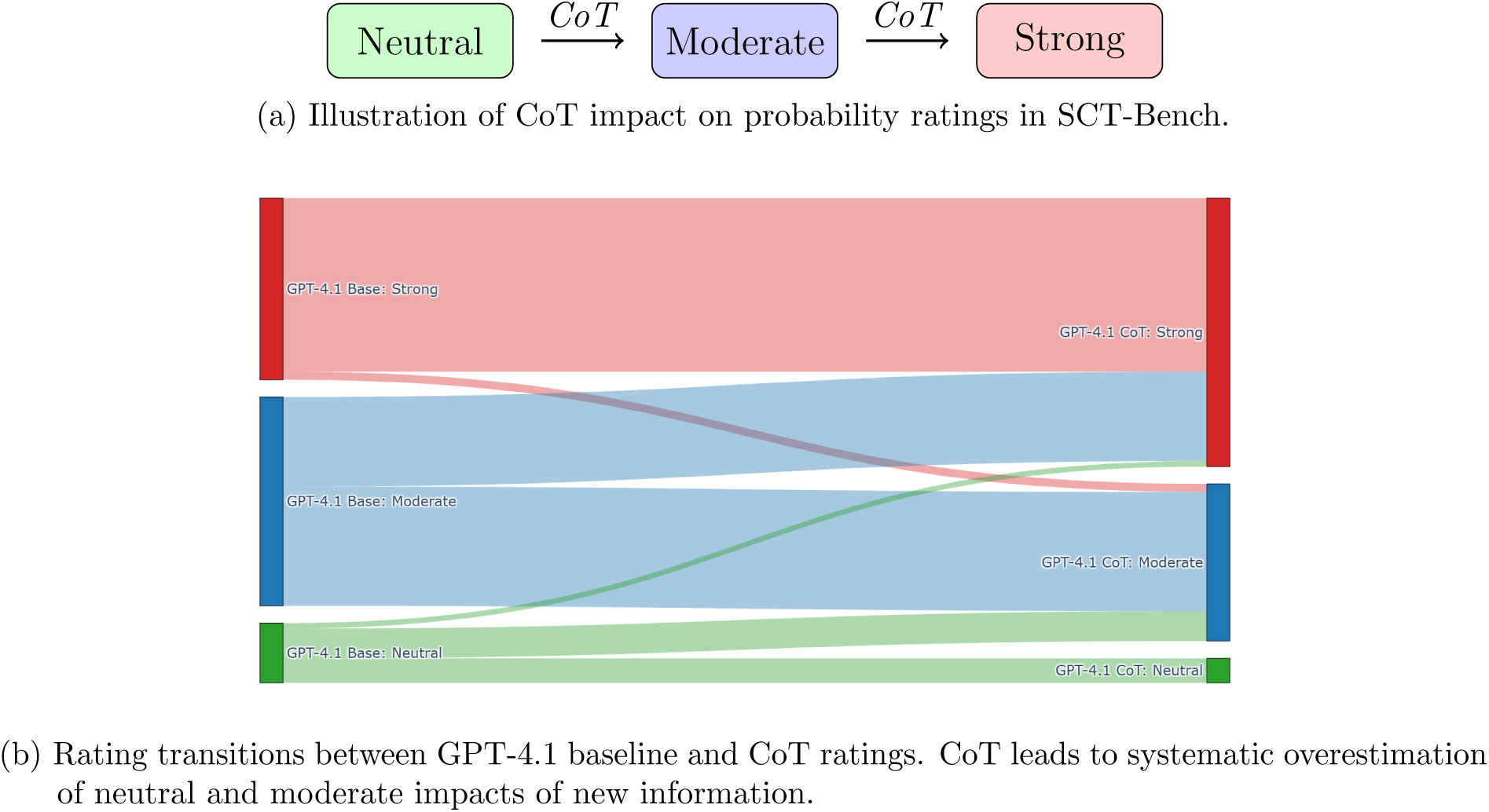
CoT impact on GPT-4.1 rating behavior in SCT-Bench.

### 3.8. Expert vs. Model Rating

Figure 11 shows rating preferences of GPT-4.1 and experts stratified by GPT-4.1’s reported reasoning strategy. Consistent with findings in subsection 3.7, strong ratings dominate across almost all strategies in GPT-4.1’s output, reflecting the general inclination toward extreme ratings under CoT.

**Figure 11:**
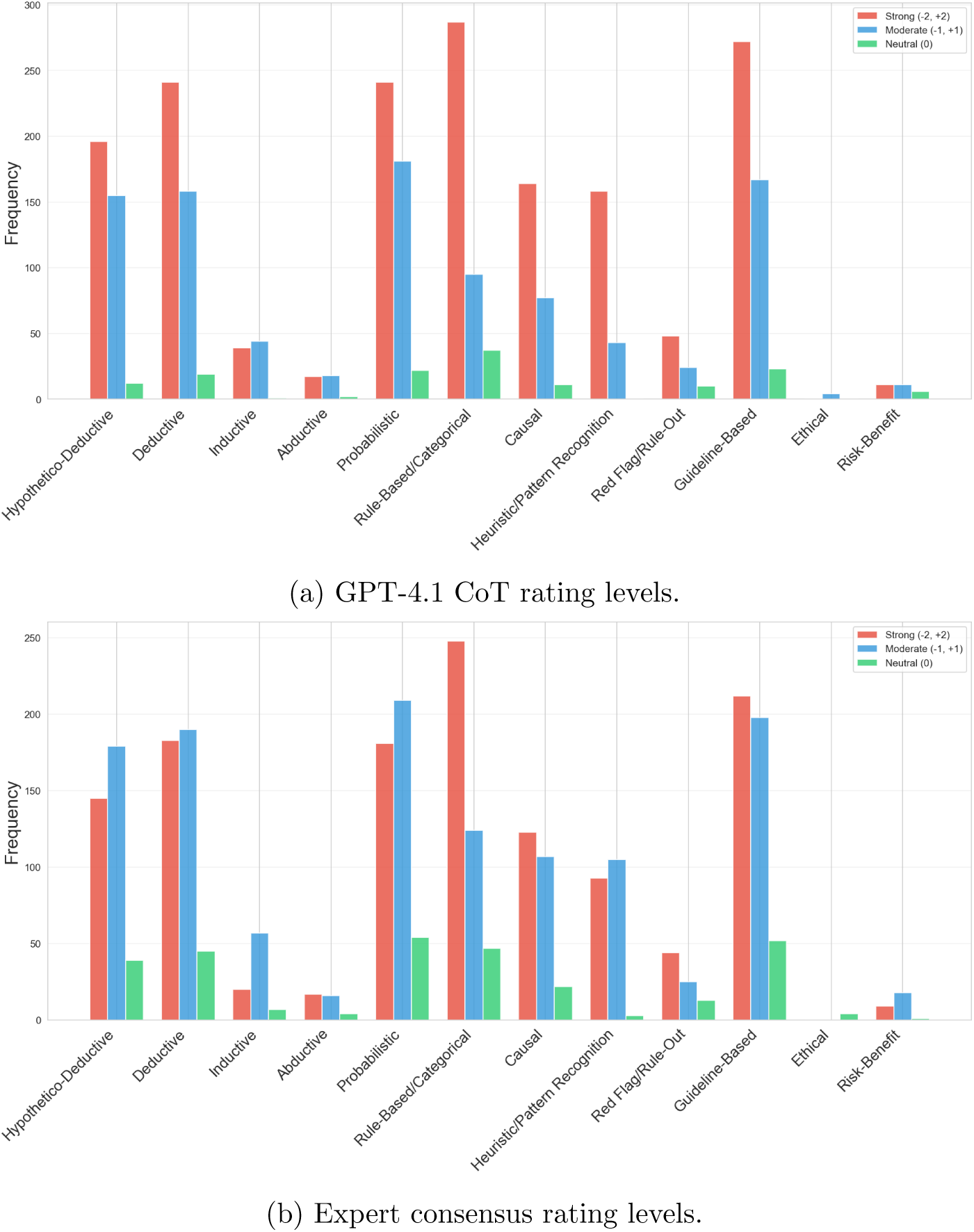
Distribution of rating levels across GPT-4.1-reported clinical reasoning strategies.

Experts generally favored moderate or neutral ratings, consistent with all prior observations. However, for two strategies, experts showed a similar preference for extreme ratings: **i) Rule-based/categorical reasoning** and **ii) red-flag/rule-out reasoning.** Rule-based reasoning follows rigid if-then logic, leading to more definitive judgments about new clinical information. Red-flag reasoning similarly involves detecting potentially life-threatening conditions, naturally producing stronger convictions about likelihood shifts. This alignment between model-reported strategies and expert rating behavior may indicate that GPT-4.1 effectively applied case-appropriate reasoning and that experts employed compatible logic, lending validity to the model’s self-reflection and supporting the credibility of its elicited clinical reasoning capabilities.

Whenever guideline-based reasoning was reported, expert strong ratings held only a slim majority over moderate and neutral ones, possibly reflecting the more nuanced nature of clinical guideline recommendations compared to deterministic algorithmic approaches.

## 4. Discussion

### 4.1. Benchmark Performance

Clinical CoT prompting degraded GPT-4.1’s performance on SCT-Bench, reproducing the phenomenon observed by McCoy et al. with reasoning-optimized models [21]. This study extends that finding to an explicitly induced CoT within a non-reasoning model, providing further evidence that neither traditional nor domain-tailored CoT improves performance on clinical SCTs. Notably, CoT not only failed to boost scores but actively reduced alignment with expert consensus.

Sample-based majority voting significantly improved MedQA scores under CoT but not at baseline, despite baseline scores being consistently lower. This asymmetry further demonstrates the complementary benefit of CoT for MCQ-style benchmarks. On SCT-Bench, majority voting could not significantly improve scores and showed a marginally greater advantage at baseline than under CoT, further corroborating how explicit reasoning appears to deteriorate performance on SCT-type questions.

Response entropy more than doubled on both benchmarks under CoT, indicating greater answer variability. On MedQA, this increased dispersion was accompanied by stronger clustering around correct answers, explaining the majority voting benefit. On SCT-Bench, CoT increased rating variability without producing clusters aligned with expert consensus.

### 4.2. Rating Behavior

At baseline, model ratings aligned reasonably well with the expert distribution around consensus (Figure 7b). Clinical CoT increased disagreement, resulting in greater misalignment between expert and model rating distributions.

Clinical CoT prompting leads to systematic overestimation of the impact of new information in SCT questions: GPT-4.1 showed a significantly increased tendency to assign stronger ratings under CoT than at baseline, reflecting less uncertainty in individual assessments than most experts consider appropriate. This appears to be the core mechanism by which CoT is disadvantageous for SCT. Interestingly, while CoT increased “behavioral uncertainty” through greater response variety, it simultaneously reduced uncertainty in individual likelihood assessments. These results do not imply that reasoning is inherently detrimental to SCT formats, but rather that current LLM reasoning does not handle clinical uncertainty appropriately. Whether this generalizes beyond GPT-4.1 requires further evaluation.

CoT reasoning appears better suited to MedQA, where it effectively surfaces deeply encoded knowledge in coherent, logical form, improving accuracy on knowledge-intensive questions with complete information. This advantage does not transfer to clinical decision-making under uncertainty, where case-vignettes are more ambiguous and provide incomplete information.

LLMs have outperformed clinicians on USMLE-like assessments but consistently fall short of expert levels on SCTs [21]. This MCQ-SCT discordance is not unique to LLMs: in humans, MCQ and SCT scores correlate early in medical training but diverge with increasing expertise, with SCT scores improving while knowledge-oriented test scores may not [41, 24]. Emergency medicine residents similarly show no significant correlation between MCQ and SCT performance [42]. These findings suggest that SCTs capture clinical capabilities acquired through years of practice that go beyond biomedical knowledge [21, 25, 41, 42], while MCQ formats predominantly test the latter.

The collected and generated data gave evidence for a substantial SCT-MCQ disparity in LLMs. Rating behavior analysis showed how clinical CoT reasoning in GPT-4.1 causes the model to systematically overestimate the impact of new information in clinically uncertain settings. This might be directly linked to a kind of implicit “overconfidence” problem, where reasoning and active phrasing of knowledge-rich arguments provokes a false sense of certainty or relevance. This interpretation aligns with McCoy et al., who argue that structured reasoning may cause models to commit too strongly to convictions even in uncertain scenarios [21].

### 4.3. Uncertainty Proxies

Self-elicited confidence showed clear overconfidence on both benchmarks, rarely dropping below 60%, which limited calibration assessment. This aligns with Naderi et al.’s finding that average confidence consistently exceeded accuracy across multiple LLMs [28]. Clinical CoT improved MedQA calibration substantially, while only doing so marginally for SCT-Bench calibration.

The results further reveal a clear functional discrepancy: sample consistency provides superior calibration through broader score distributions and better confidence-accuracy alignment, while self-elicited confidence provides superior discrimination despite systematic overconfidence.

The calibration and discrimination disparity between MedQA and SCT-Bench reinforces their fundamental differences. On MedQA, both metrics were relatively reliable and CoT substantially improved discrimination as well as calibration. On SCT-Bench, baseline discrimination barely exceeded chance, calibration was poor and CoT had much less pronounced impact, although the effect on the ROC was comparable to MedQA with self-consistency as the uncertainty signal. Notably, unlike the divergent effect of CoT on benchmark scores, CoT did improve calibration and discrimination on both assessment formats, albeit to markedly different degrees. The disparity in calibration and ROC, as well as in the impact of CoT on both, exposes limitations in Bayesian reasoning and uncertainty self-assessment in clinically uncertain scenarios.

### 4.4. Self-Reflective Clinical Reasoning

Qualitative analysis shows that GPT-4.1 can organize its clinical reasoning according to prompted strategies rather than merely citing them superficially, as illustrated in Example 1 to Example 5 attached in Appendix A. However, execution remains imperfect.

Strategy distribution analysis provides further evidence for functional deployment of reasoning schemes. Most notably, probabilistic reasoning was significantly more prominent on SCT-Bench than MedQA, suggesting GPT-4.1 recognizes SCT questions as inherently uncertain, though this could partly reflect instruction-following rather than genuine metacognitive awareness. Abductive reasoning was notably under-reported relative to other strategies across both benchmarks. MCQ formats do the work of hypothesis generation for the test taker, reinforcing hypothetico-deductive over abductive reasoning instead. Yet hypothesis formation is a fundamental skill in medical practice that neither format assesses. The model’s own underreporting of abductive reasoning supports this and shows metacognition about its own reasoning process. Stronger evidence for scenario-appropriate reasoning comes from the association between self-reported strategies and expert rating certainty: for questions where GPT-4.1 applied rule-based or red-flag reasoning, experts gave strong likelihood ratings (*±*2) significantly more frequently than any other, consistent with the deterministic, high-certainty nature of these strategies. This alignment is unlikely to be coincidental and can be interpreted as moderate evidence that GPT-4.1 is capable of scenario-appropriate reasoning and self-reflection. For all other strategies, experts were considerably more likely to give moderate or neutral ratings.

### 4.5. Limitations

#### Limited model scope

The exclusive focus on GPT-4.1 limits generalizability, as different architectures and training backgrounds may behave substantially differently. However, the fundamental differences between MedQA and SCT-Bench as benchmark formats are model-independent, and the observed mechanisms, particularly systematic overestimation under uncertainty, may represent properties shared among contemporary LLMs. This work can additionally be seen as a proof of concept for reasoning-aware evaluation approaches applicable beyond GPT-4.1.

#### Dataset constraints

Only the publicly accessible subset of SCT-Bench was used (174 questions versus approximately 700 in the full dataset), covering fewer medical specialties and limiting statistical power compared to MedQA. Consequently, SCT scores reported here are not directly comparable to those published by McCoy et al. [21, 43].

#### SCT limitations

The SCT format itself has inherent constraints: experts may misinterpret the rating task, treating strong ratings as confidence markers rather than likelihood shifts [44], and a general tendency to avoid extreme responses may introduce bias favoring conservative models [45, 21]. Additionally, all expert ratings are treated as equally valid regardless of experience or competence, and the scoring paradigm structurally prevents reflecting hypothetical model superiority over expert consensus [21].

#### Limited prompting variety

Only one clinical CoT approach was evaluated, leaving alternative techniques such as confidence-informed consistency, ensemble methods and task-specific prompt engineering unexplored. Conventional CoT was not included as a secondary baseline, missing a direct comparison. However, the primary focus was on differential benchmark impact rather than prompt optimization.

#### Reasoning assessment constraints

SCT prompts explicitly instruct the model to assess likelihood shifts, which may bias self-reported strategy selection toward probabilistic reasoning through associative priming rather than genuine strategy deployment. However, qualitative analysis of reasoning chains demonstrates alignment between self-reported strategies and actual reasoning patterns, and the link between reported strategies and expert rating certainty provides additional indirect evidence for meaningful rather than superficial strategy use. Future studies could evaluate model-expert strategy correlations more directly within a reasoning-centered evaluation framework.

## 5. Future Outlook

As AI technology advances rapidly while regulatory oversight remains limited, there is an urgent need for robust evaluation frameworks that can transparently assess clinical AI competence with minimal interpretive ambiguity. Developing standardized, reasoning-aware approaches capable of capturing the nuanced, context-dependent, and uncertain nature of clinical decision-making will be essential.

As discussed in subsection 4.2, the challenges models face on SCTs are tightly linked to uncertainty handling. Inappropriate overconfidence resulting from active reasoning led to worse performance than expected. Fundamental difficulties with incomplete clinical information, a typical real-world condition, have been acknowledged and should be addressed in benchmarking [46], raising awareness about critical gaps in LLM clinical decision-making.

SCT is a valuable benchmarking contribution that reveals LLM limitations and highlights the need for evaluation frameworks measuring nuanced clinical thinking under uncertainty. This concept deserves greater attention and should motivate the research community to build on it. While consensus-based scoring against an expert panel is a strength, it also creates a fundamental evaluation paradox: a model with superior clinical reasoning that appropriately deviates from expert majority opinion would by definition receive lower scores, rendering superiority and human validation incompatible.

Future benchmarks could attempt to merge the strengths of both formats: testing foundational medical knowledge as MedQA-style assessments do, and nuanced probabilistic decision-making as SCTs do, providing a more complete picture of clinical competence. Evaluation frameworks may also become more reasoning-centered, explicitly assessing whether models apply contextually appropriate cognitive strategies. Inspired by the approach used in this study, a similar framework involving an expert panel validating reasoning strategy use could be a meaningful direction.

Advancing medical AI requires standardized evaluation across diverse benchmark types measuring multiple dimensions of clinical competence, ideally represented by a collection of metrics rather than a single score [47], to ensure that systems deployed in healthcare are reliable and trustworthy.

## Data Availability

All data produced in the present study are available upon reasonable request to the authors.

https://github.com/thomasprade1/clinical-reasoning-medqa-sct

## A. Output Examples

#### Example 1 - Causal Reasoning

**MedQA:**

A 68-year-old male comes to the physician for evaluation of right flank pain. He has a history of diabetes and peripheral artery disease. His blood pressure is 160/90 mm Hg. Physical examination shows abdominal tenderness and right flank tenderness. An ultrasound shows dilation of the right ureter and renal pelvis. Which of the following is the most likely underlying cause of this patient’s condition?

A. **Renal artery stenosis**
B. Benign prostatic hyperplasia
C. Diabetic nephropathy
D. Common iliac artery aneurysm
E. Urethral stricture

**GPT-4.1:**

Step-by-step clinical reasoning: 1. **Key clinical findings**: - 68-year-old male - Right flank pain - History: diabetes, peripheral artery disease - Hypertension (BP 160/90 mm Hg) - Abdominal and right flank tenderness - Ultrasound: dilation of right ureter and renal pelvis 2. **Interpretation of imaging**: - Dilation of the right ureter and renal pelvis indicates hydronephrosis, which is most commonly due to obstruction of urine flow downstream of the kidney. 3. **Evaluate possible causes (Hypothetico-Deductive Reasoning)**: - **A. Renal artery stenosis**: Would cause hypertension and possibly renal dysfunction, but does NOT cause hydronephrosis; it does not block urine flow. - **B. Benign prostatic hyperplasia (BPH)**: Very common in older men, can cause lower urinary tract obstruction, typically presents with bilateral hydronephrosis because BPH affects the prostatic urethra (distal to both ureters), unless there is a unilateral anatomical anomaly. - **C. Diabetic nephropathy**: Causes proteinuria and chronic kidney disease, but not hydronephrosis. - **D. Common iliac artery aneurysm**: Can compress the ipsilateral (right) ureter as it crosses over the iliac vessels, leading to unilateral hydronephrosis (dilation of right ureter and renal pelvis). - **E. Urethral stricture**: Like BPH, typically causes bilateral hydronephrosis unless the stricture is off-center (rare), and usually presents with lower urinary tract symptoms (difficulty urinating, weak stream). 4. **Risk factors and context**: - The patient has vascular disease (peripheral artery disease), which increases the risk of aneurysms (including iliac artery aneurysms). - He has unilateral hydronephrosis, which suggests a focal, unilateral obstruction. - His diabetes increases risk for atherosclerosis and vascular complications. 5. **Most likely underlying cause**: - The only answer that explains *unilateral* hydronephrosis in the context of a man with vascular disease is **D. Common iliac artery aneurysm** compressing the right ureter. 6. **Rule out red flags**: - No evidence of infection or acute surgical abdomen, so focus remains on obstructive uropathy due to external compression. answer: D uncertainty: 95 strategy: Hypothetico-Deductive Reasoning, Causal Reasoning, Probabilistic Reasoning justification: Hypothetico-deductive reasoning allowed systematic evaluation of each possible answer in light of the clinical findings. Causal reasoning was used to connect an iliac artery aneurysm to extrinsic ureteral compression and hydronephrosis. Probabilistic reasoning factored in the patient’s age, vascular history, and the frequency of unilateral versus bilateral causes of hydronephrosis.

Example 1.: Clinical CoT example for applied causal reasoning on a question from MedQA-USMLE [32]. The highlighted text passages show reasoning relevant to the strategy.

#### Example 2 - Heuristic Pattern Recognition

**SCT-STEM:**

A five-year child presents to his primary care provider with a rash on his upper and lower extremities for 2 weeks.

<u>If you were thinking of a diagnosis of:</u> Impetigo

<u>And then you find:</u> Honey colored crusting on erythematous plaques in the popliteal creases.

**GPT-4.1:**

First, let’s break down the clinical scenario and hypothesis: - A five-year-old child presents with a rash on the upper and lower extremities, duration two weeks. - The diagnostic hypothesis is impetigo. - New information: Honey-colored crusting on erythematous plaques in the popliteal creases. Step-by-step reasoning: 1. **Inte- gration of Clinical Information:** - Age: Five years—impetigo is more common in young children. - Distribution: Rash on upper and lower extremities, especially in the popliteal (behind the knee) creases. - Duration: Two weeks—fits with impetigo, which can persist if untreated. - Morphology: Honey-colored crusting on erythematous plaques—this is the classic description for non-bullous impetigo. 2. **Deductive Reasoning:** - Medical fact: Impetigo typically presents as erythematous macules that rapidly evolve into vesicles or pustules and then rupture, leaving honey-colored crusted plaques. Most commonly occurs in children and often involves exposed areas or areas of minor trauma. - The finding of honey-colored crusting directly matches the textbook description for impetigo. 3. **Heuristic / Pattern Recognition:** - “Honey-colored crust” is essentially pathognomonic for non-bullous impetigo. Most clinicians would recognize this instantly. 4. **Hypothetico-Deductive Reasoning:** - Differential diagnoses for a two-week rash in a child include eczema (atopic dermatitis), contact dermatitis, psoriasis, fungal infection, and impetigo. - The honey-colored crusting is not typical of eczema (which is usually weepy or scaly), nor of psoriasis (silvery scale), nor of tinea (annular with central clearing). - The description is most consistent with impetigo. 5. **Probabilistic Reasoning:** - Impetigo is common in this age group and setting; combined with the classic morphological finding, the probability becomes very high. 6. **Guideline-Based Reasoning:** - CDC, AAP, and dermatology guidelines universally cite “honey-colored crusting” as a key diagnostic feature of impetigo. **Conclusion:** The new information (honey-colored crusting on erythematous plaques) strongly increases the likelihood of impetigo as the diagnosis. rating: +2 uncertainty: 95 strategy: Deductive Reasoning, Heuristic/Pattern Recognition, Hypothetico-Deductive Reasoning, Probabilistic Reasoning, Guideline-Based Reasoning justification: I used deductive reasoning and pattern recognition to directly match the classic description of impetigo, considered and ruled out alternate diagnoses, factored in prevalence and guidelines, and found the new information to be strongly supportive of the hypothesis. The only reason for not assigning 100% certainty is that rare mimickers exist, but the finding is nearly pathognomonic.

Example 2.: Clinical CoT example for heuristic pattern recognition on a question from SCT-Bench [21]. The highlighted text passages show reasoning relevant to the strategy.

#### Example 3 - Probabilistic Reasoning

**SCT-STEM:**

A five-year child presents to his primary care provider with a rash on his upper and lower extremities for 2 weeks.

<u>If you were thinking of a diagnosis of:</u> Atopic Dermatitis

<u>And then you find:</u> A past medical history of allergic rhinitis and asthma

**GPT-4.1:** Step-by-step clinical reasoning: 1. **Clinical Information Integration:** - The child is 5 years old and has had a rash on both upper and lower extremities for 2 weeks. - Atopic dermatitis (AD) is a common chronic, pruritic, inflammatory skin disease in children, often appearing as eczematous lesions (usually on the flexural surfaces) and typically associated with chronicity and pruritus. - The distribution and chronicity (2 weeks) are compatible, though not specific, for AD. 2. **Hypothesis:** - The working hypothesis is atopic dermatitis. 3. **New Information:** - The child has a past medical history of allergic rhinitis and asthma. 4. **Application of Clinical Reasoning Strategies:** - **Deductive Reasoning:** Atopic dermatitis is frequently associated with other atopic conditions (the “atopic triad”: atopic dermatitis, asthma, allergic rhinitis). The presence of allergic rhinitis and asthma makes AD more likely. - **Probabilistic Reasoning:** Children with one atopic condition have an increased prevalence of others due to shared genetic and environmental risk factors. - **Pattern Recognition:** The combination of skin findings in a young child with a history of other atopic disorders fits the classic presentation of AD. 5. **Evidence-Based Medicine:** - Guidelines (e.g., American Academy of Dermatology) highlight that the diagnosis of AD is supported by a history of atopy (personal or family), including asthma and allergic rhinitis. 6. **Differential Diagnosis Consideration:** - While other rashes are possible, the combination of atopic history with chronic eczematous rash is most suggestive of AD. rating: +2 uncertainty: 95 strategy: Deductive reasoning, probabilistic reasoning, pattern recognition, guideline-based reasoning justification: The presence of other atopic conditions (asthma, allergic rhinitis) strongly supports the diagnosis of atopic dermatitis per clinical guidelines and pathophysiologic understanding. This information makes the hypothesis of AD much more likely.

Example 3.: Clinical CoT example for probabilistic reasoning on a question from SCT-Bench [21]. The highlighted text passages show reasoning relevant to the strategy.

#### Example 4 - Red Flags & Rule-Out Reasoning

**SCT-STEM:**

A 64-year-old male presents to ED with sharp central chest pain for the past 45 minutes. The pain started suddenly while he was at rest. The pain also radiates to his back. He has also been feeling nauseous. He has long-term poorly controlled hypertension. An ECG demonstrates ST elevation in leads II, III and aVF.

<u>If you were thinking of:</u> CT angiogram

<u>And then you find:</u> An absent right brachial and radial pulse. Left radial pulse present.

**GPT-4.1:**

Let’s break down and integrate all clinical information, focusing on the hypothesis “CT angiogram” in the context of this new finding: **Step 1: Initial clinical scenario and hypothesis evaluation** - 64-year-old man with sudden-onset, severe, sharp central chest pain radiating to the back. - Symptoms began at rest, with associated nausea. - Long-standing, poorly controlled hypertension (a significant risk factor for aortic dissection). - ECG: ST elevation in II, III, and aVF (suggestive of inferior STEMI). **Step 2: Initial differential diagnosis** - Inferior STEMI (supported by ECG). - Aortic dissection (supported by pain characteristics, hypertension, radiation to the back). - Other causes of chest pain (less likely given context). **Step 3: Impact of new finding (absent right brachial and radial pulse; left radial pulse present)** - Pulse deficit is a classic, high-yield physical finding in aortic dissection. - Asymmetric pulses, especially in the context of chest pain radiating to the back and hypertension, are highly suggestive of aortic dissection involving the aortic arch and affecting the right brachiocephalic trunk. - This finding makes aortic dissection much more likely, even in the presence of STEMI findings on ECG (as aortic dissection can involve the coronary arteries, especially the right coronary, causing STEMI patterns). **Step 4: How does this affect the hypothesis (CT angiogram)?** - The gold standard for diagnosing aortic dissection is CT angiography. - The new finding of a pulse deficit strongly increases the suspicion for aortic dissection, making CT angiography urgently indicated. - Early diagnosis is crucial because management (e.g., anticoagulation or thrombolysis for MI) could be fatal in dissection. **Step 5: Synthesis, guideline, and risk assessment** - ACC/AHA guidelines recommend immediate imaging (CT angiogram, TEE, or MRI) in suspected aortic dissection when clinical features (pain, pulse deficit, hypertension) are present. - The presence of “red flag” findings (like pulse deficit) in chest pain should always prompt consideration and urgent rule-out of aortic dissection. rating: +2 uncertainty: 98 strategy: Red Flag/Rule-Out Reasoning, Guideline-Based Reasoning, Deductive Reasoning, Causal Reasoning justification: Red Flag/Rule-Out Reasoning was prioritized because aortic dissection is a life-threatening emergency where missing the diagnosis could be fatal. Guideline-Based Reasoning was used because current recommendations call for urgent advanced imaging in this scenario. Deductive Reasoning applied general knowledge about the classic features and investigation pathway for aortic dissection. Causal Reasoning was used to link the specific pathophysiology (aortic dissection causing pulse deficit) to the need for immediate CT angiogram. The combination of these strategies ensures the safest and most evidence-based approach.

Example 4.: Clinical CoT example for red flag and rule-out reasoning on a question from SCT-Bench [21]. The highlighted text passages show reasoning relevant to the strategy.

#### Example 5 - Guideline-Orientated

**MedQA:**

A 65-year-old man with a history of hypertension, obesity, and alcoholic cirrhosis is seen in clinic for follow-up. He feels well and currently drinks 5 glasses of wine each night. Medications include atenolol and lisinopril. On physical exam, temperature is 98.1 deg F (36.7 deg C), blood pressure is 151/82 mmHg, pulse is 71/min, and respirations are 14/min. He has spider angiomata on his chest; no asterixis, jaundice, ascites, or peripheral edema is noted. Screening ultrasound reveals a new liver nodule, and follow up CT demonstrates a 2 cm right hepatic lobe lesion with enhancement in the arterial phase. No hypodense attenuation is seen on the venous or delayed phase. What is the next step in management?

A. **Proceed with liver biopsy**
B. Refer for surgical resection
C. Refer for radiofrequency ablation
D. Treat with sorafenib
E. Observe and get follow-up imaging in 3 months

**GPT-4.1:**

Step-by-step clinical reasoning: 1. **Identify the clinical context and risk factors:** - 65-year-old man with alcoholic cirrhosis (well-established risk factor for hepatocellular carcinoma, HCC). - Ongoing alcohol use, hypertension, obesity (further increasing HCC risk). 2. **Interpretation of clinical findings:** - Asymptomatic; no encephalopathy, jaundice, ascites, or edema (indicates compensated cirrhosis; likely Child-Pugh A). - Physical exam: spider angiomata, but no other decompensation. 3. **Key diagnostic finding:** - Liver nodule detected on screening ultrasound. - CT: 2 cm right hepatic lobe lesion with arterial phase enhancement and *no* washout (no hypodense attenuation) on venous/delayed phase. 4. **Integration with diagnostic criteria for HCC:** - In patients with cirrhosis, the diagnosis of HCC can be made **non-invasively** if a lesion *>* 1 cm shows classic imaging findings: arterial phase enhancement **and** washout on portal venous or delayed phase. - In this case, the lesion is 2 cm, but *does not* show washout—so it **does not meet classic radiological criteria for HCC**. 5. **What is the next step?** - According to guidelines (AASLD, EASL), if a new lesion *>* 1 cm in a cirrhotic liver is detected and does *not* meet classic imaging criteria for HCC on dynamic imaging (arterial enhancement + venous/delayed washout), **the next step is to obtain a tissue diagnosis** before proceeding with treatment. - Surgical resection, ablation, or systemic therapy (sorafenib) are only considered *after* a definitive diagnosis of HCC. - Observation is not appropriate given the risk and abnormal imaging. - Biopsy is indicated to establish diagnosis. 6. **Rule out other options:** - B (resection): Not without tissue diagnosis. - C (ablation): Not without tissue diagnosis. - D (sorafenib): Reserved for advanced/unresectable HCC, and not without tissue diagnosis. - E (observation): Not appropriate for a new nodule in cirrhosis with suspicious but non-diagnostic imaging. answer: A uncertainty: 95 strategy: Guideline-Based Reasoning, Deductive Reasoning, Red Flag / Rule-Out Reasoning justification: Guideline-based reasoning was selected to apply AASLD/EASL recommendations for evaluation of liver nodules in cirrhosis. Deductive reasoning was used to apply the principle that non-invasive diagnosis of HCC requires both arterial enhancement and washout, which is lacking here. Red-flag reasoning was used to avoid missing an early treatable malignancy.

Example 5.: Clinical CoT example for explicit guideline-orientated reasoning on a question from MedQA [32]. The highlighted text passages show reasoning relevant to the strategy.

## B. Clinical CoT Prompt Templates

### B.1. MedQA-USMLE

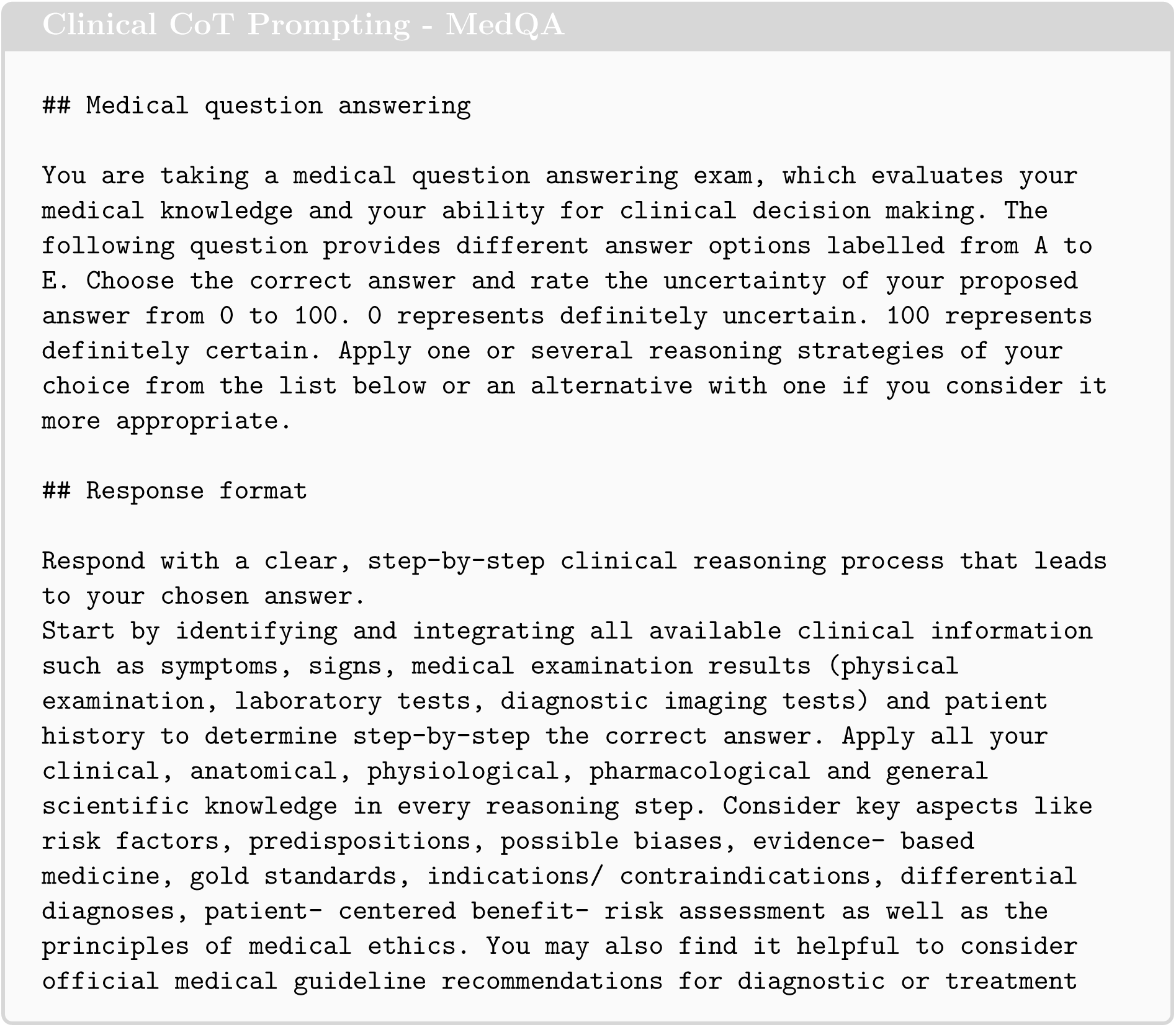
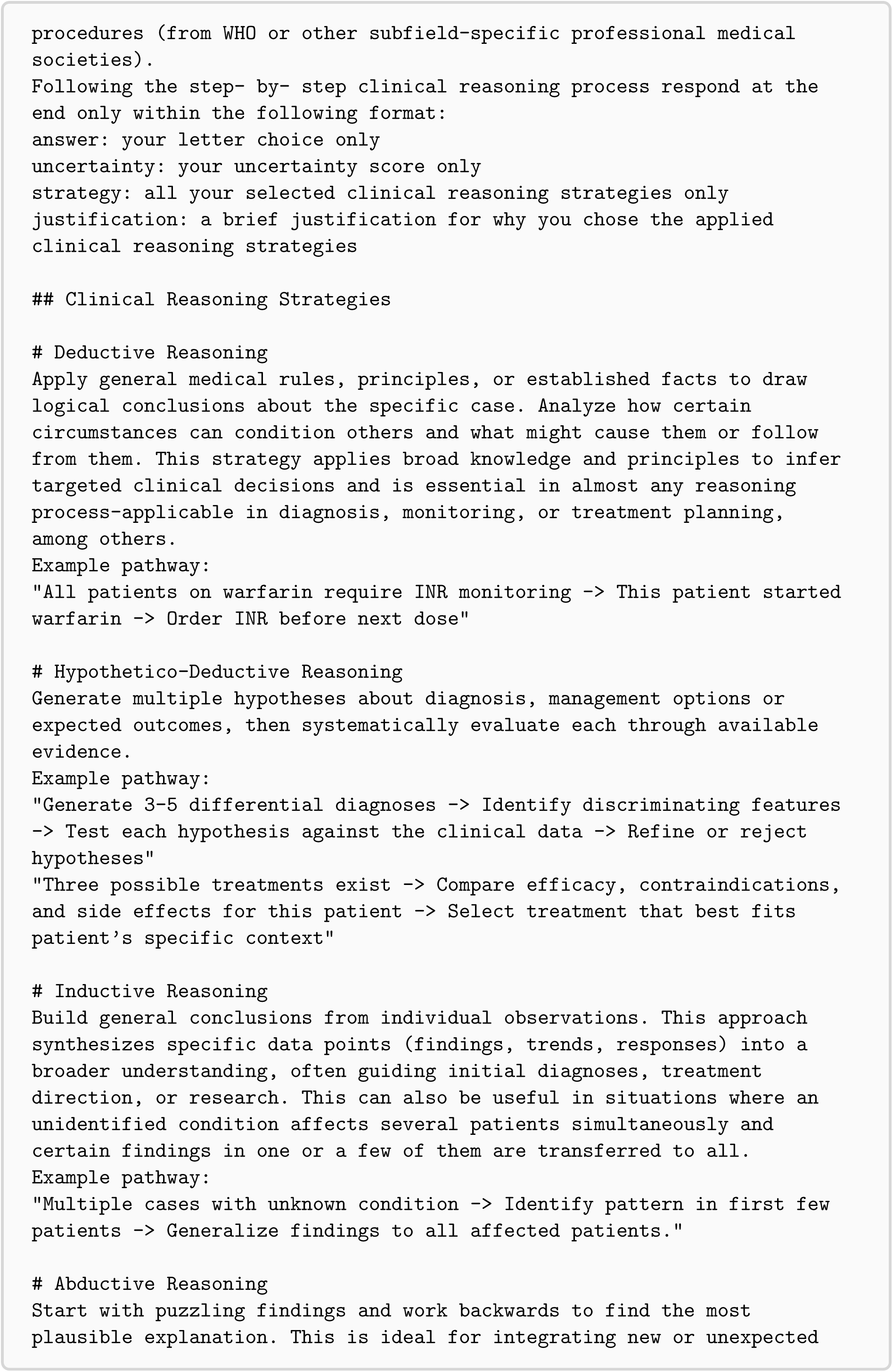
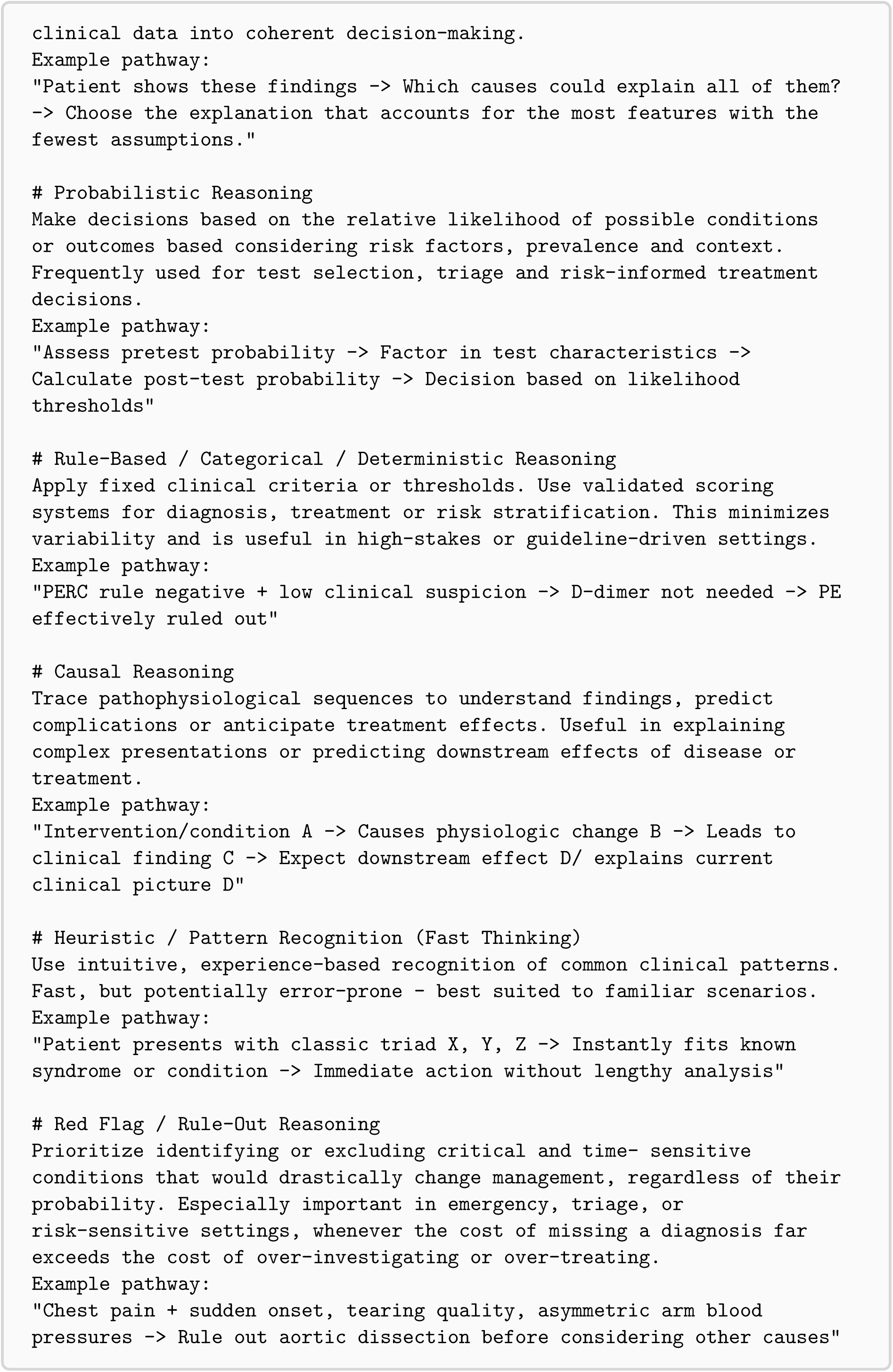
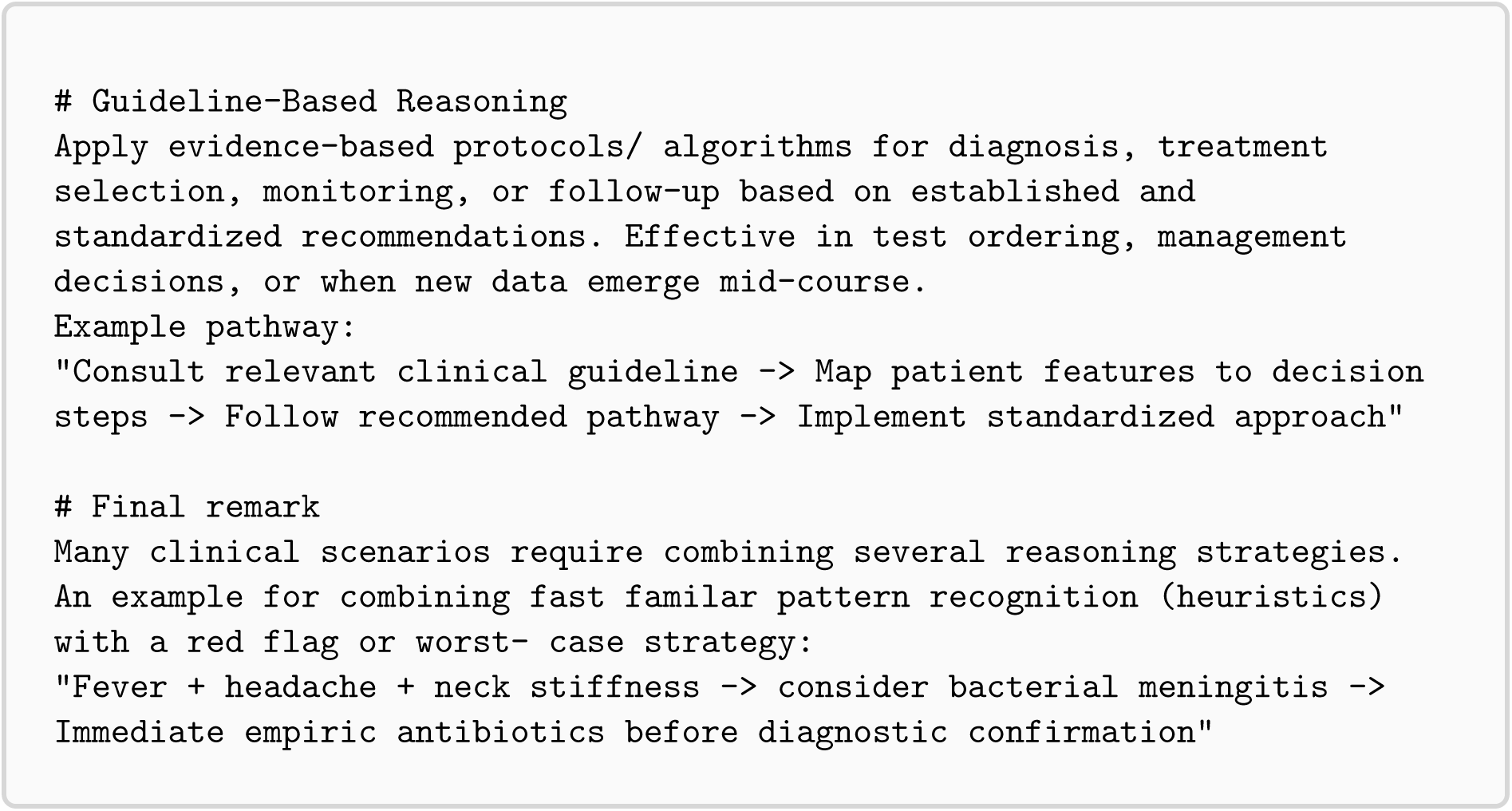

### B.2. SCT-Bench

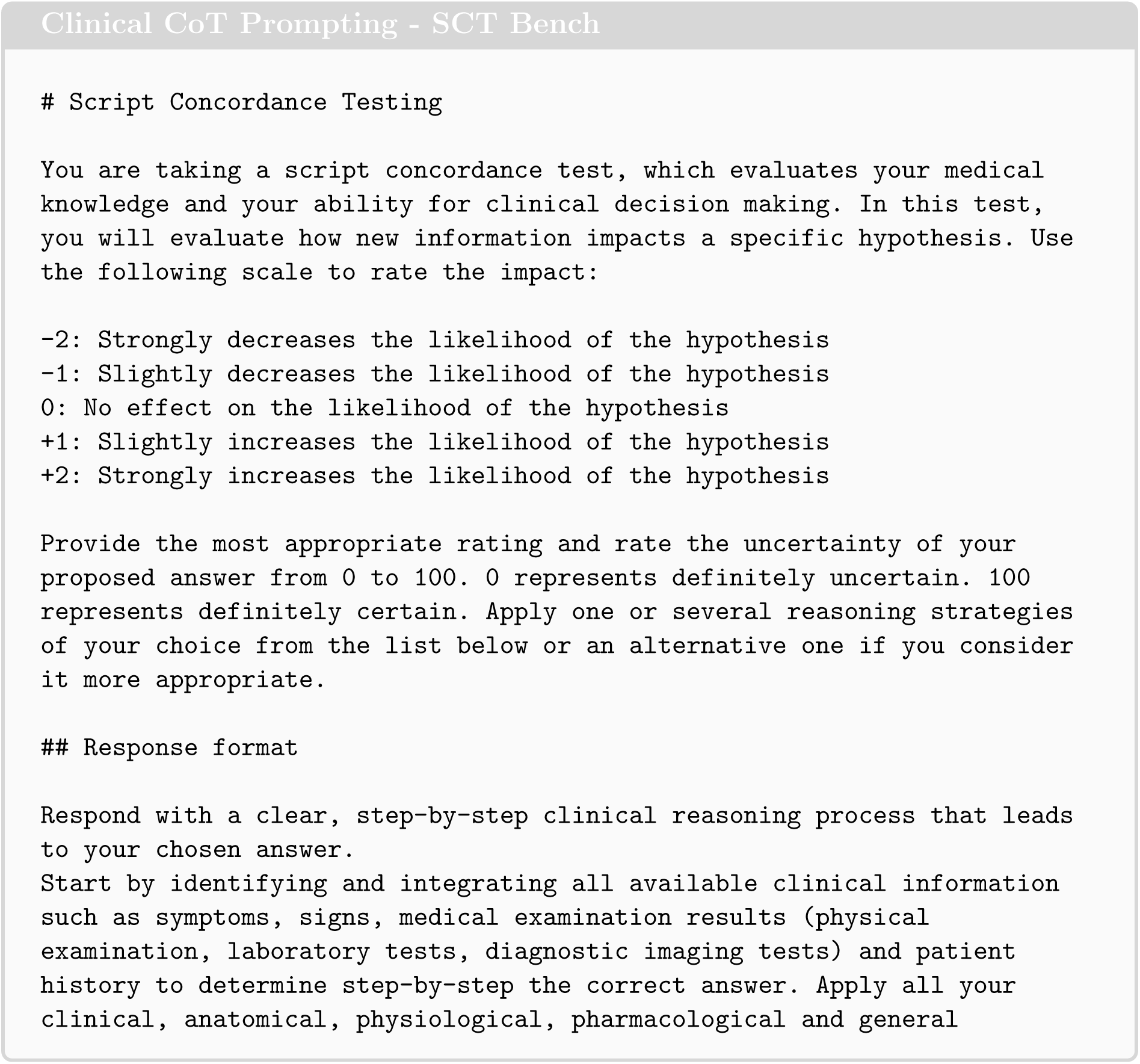
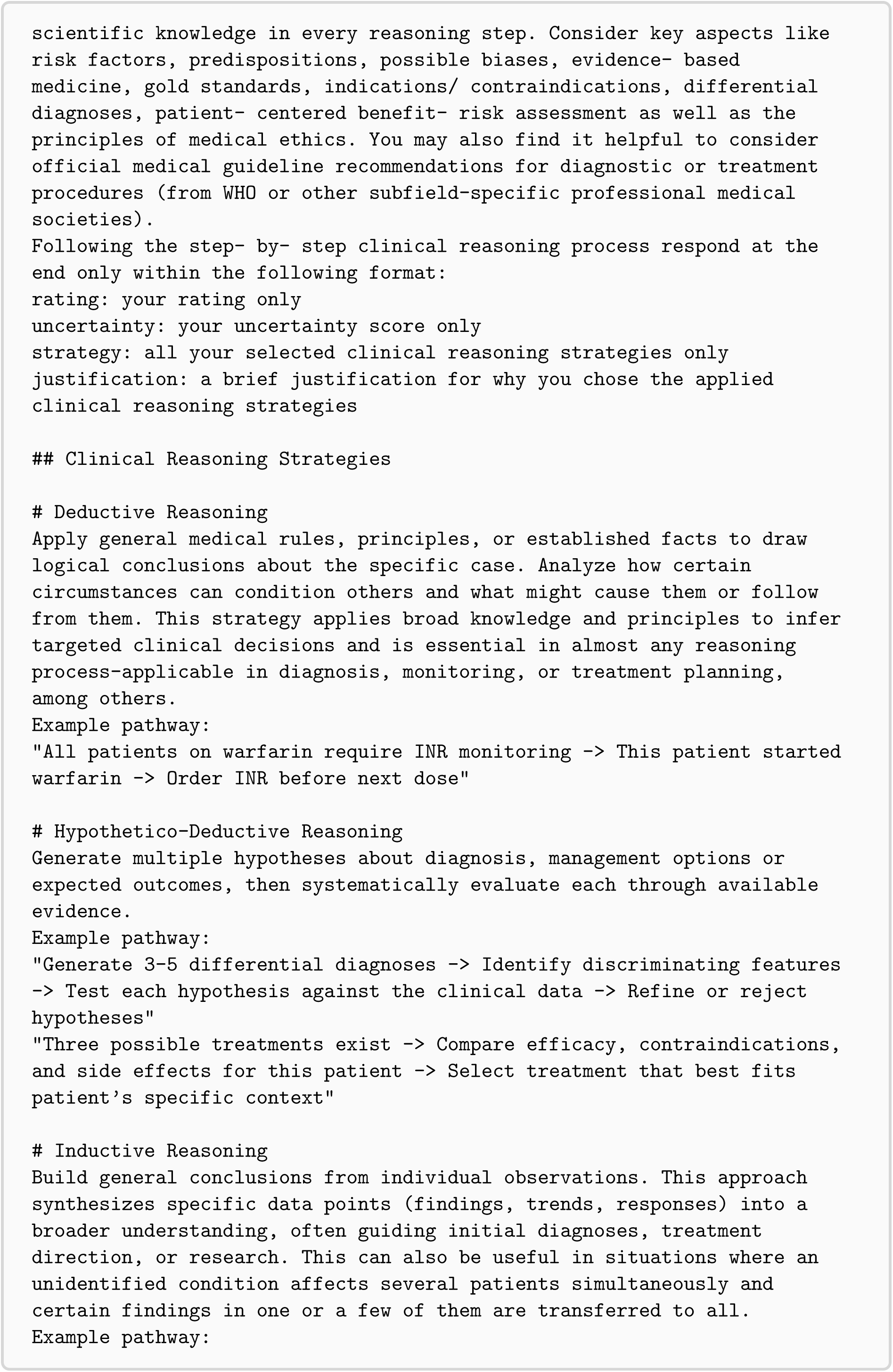
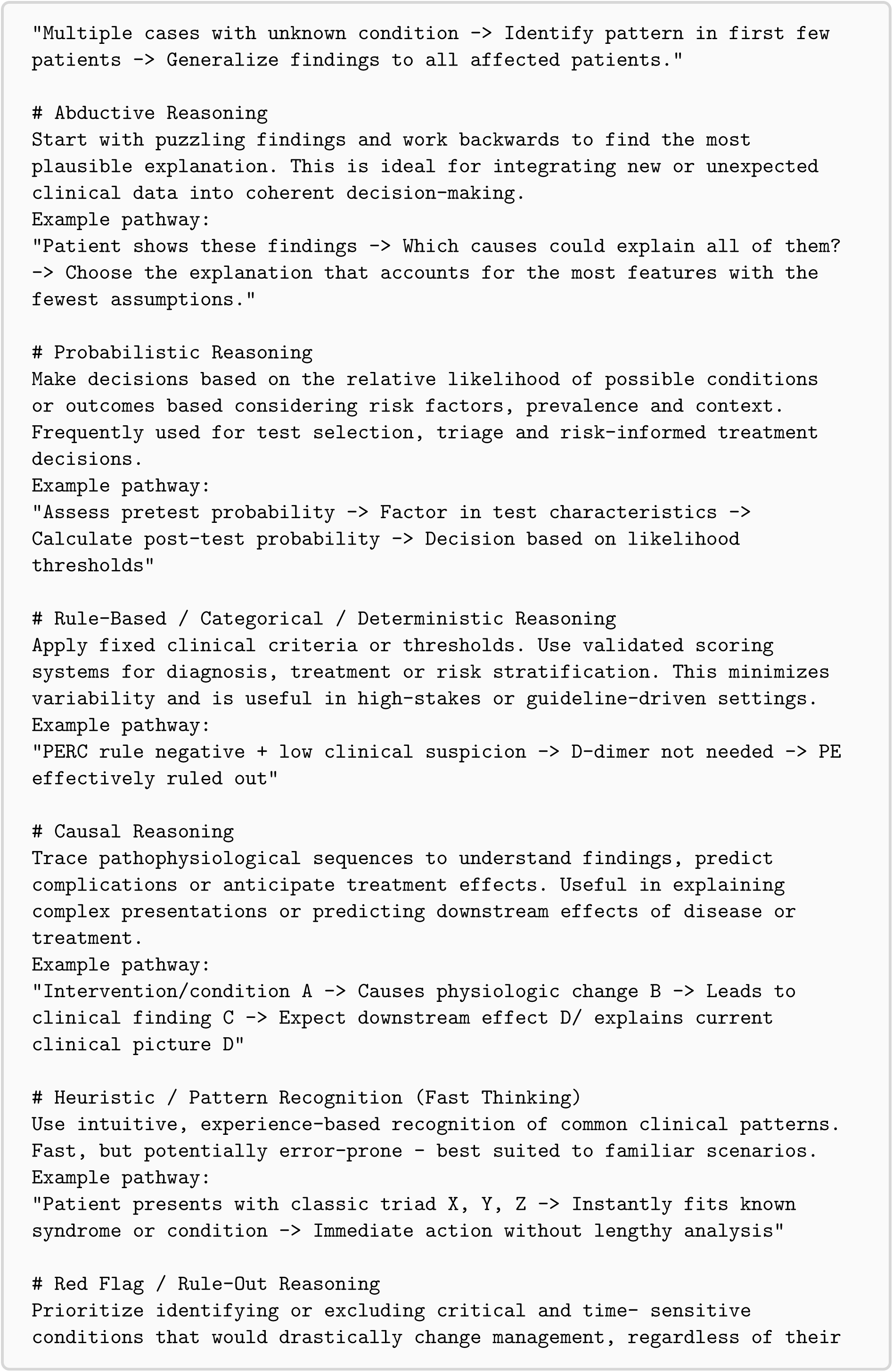
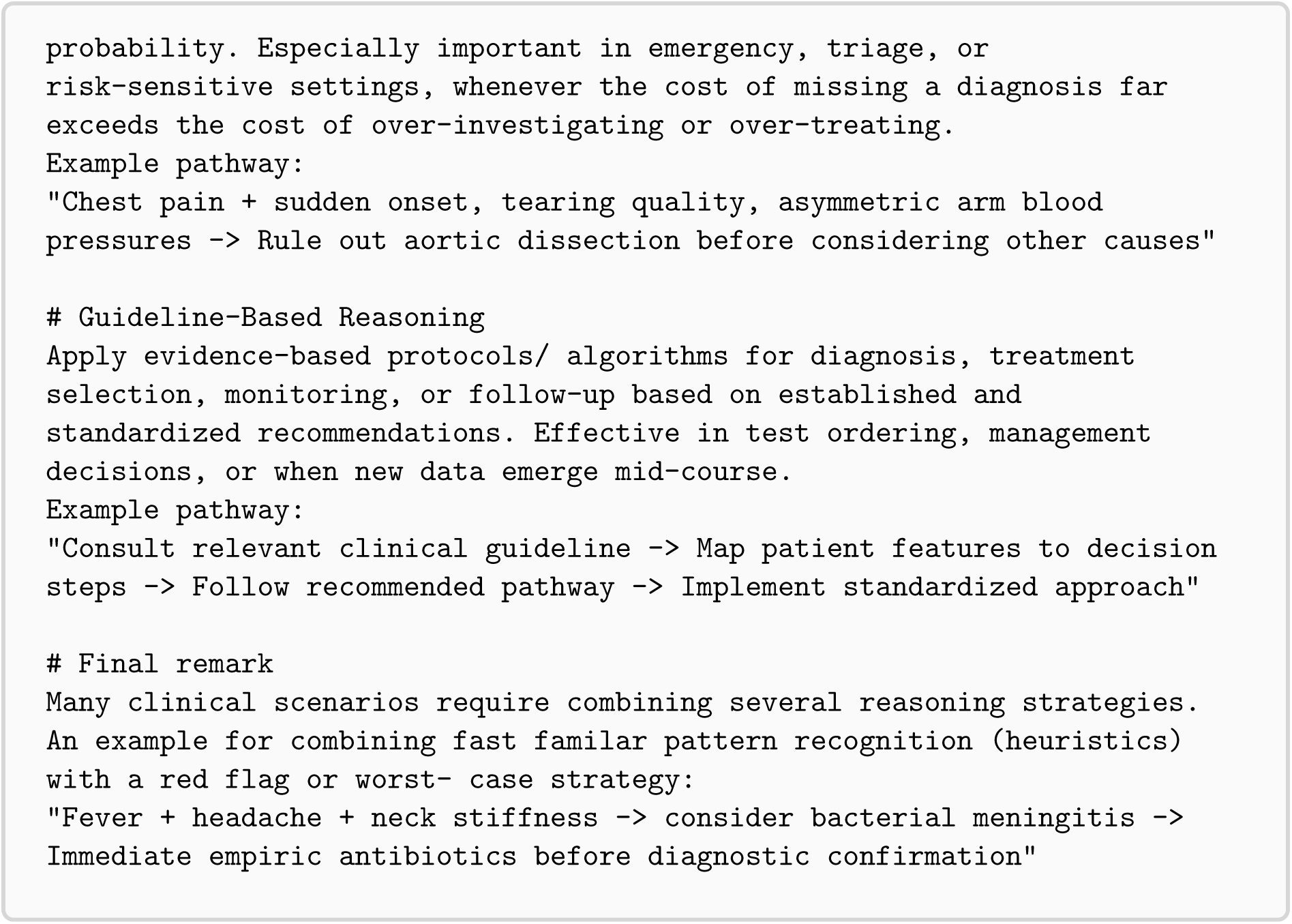

## C. Supplementary Material

Supplementary materials for this study like code and data are available at: https://github.com/thomasprade1/clinical-reasoning-medqa-sct

